# The impact of the coronavirus disease 2019 (COVID-19) outbreak on cancer practice in Japan: using an administrative database

**DOI:** 10.1101/2020.11.18.20233700

**Authors:** Hisashi Itoshima, Yuka Asami, Jung-ho Shin, Daisuke Takada, Tetsuji Morishita, Seiko Bun, Takuya Okuno, Susumu Kunisawa, Yuichi Imanaka

## Abstract

**Background:** Recent researches reported the impact of the coronavirus disease 2019 (COVID – 19) pandemic on the clinical practice of specific type cancers. The aim of this study was to reveal the impact of the COVID-19 outbreak on the clinical practice of various cancers.

**Methods:** We included hospitalized patients aged 18 years or older diagnosed between July 2018 and June 2020 with one of the top 12 most common cancers in Japan (colon/rectum, lung, gastric, breast, bladder & urinary tract, pancreas, non-Hodgkin lymphoma, liver, prostate, esophagus, uterus, and gallbladder & biliary tract) using Diagnostic Procedure Combination data, an administrative database in Japan. The intervention was defined April 2020 based on a declaration of emergency from Japanese government. The change volume of number of monthly admissions with each cancer was tested by interrupted time series (ITS) analysis, and monthly cases with radical surgery or chemotherapy for each cancer were descripted.

**Results:** 403,344 cases were included during the study period. The most common cancer was colon/rectum (20.5%), followed by lung (17.5%). In almost cancer cases, the number of admissions decreased in May 2020. In particular, colorectal, lung, gastric, breast, uterine, or esophageal cancer cases decreased by over 10%. The number of admissions with surgery or chemotherapy decreased in colorectal, lung, gastric, breast, uterine, or esophageal cancer. ITS analysis indicated that cases with gastric or esophageal cancer were affected more than other type of cancer.

**Conclusions:** The COVID-19 outbreak has a negative impact on the number of admission cases with cancer; the magnitude of impact varied by cancer diagnosis.

## Introduction

The outbreak of severe acute respiratory syndrome coronavirus 2 (SARS-Cov-2) is affecting a wide range of clinical practices. Since the outbreak in December 2019, more than 30 million cases of coronavirus disease 2019 (COVID-19) have been reported worldwide, including more than 959,000 deaths [1]. The symptoms of COVID-19 vary from fever and common cold symptoms to acute respiratory distress syndrome, septic shock, and death [2].

In the field of oncology, many academic societies, educational institutions, and government agencies have announced recommendations during COVID-19 pandemic [3-6]. The European Society for Medical Oncology (ESMO) has divided its treatment priorities into three phases (High, Medium, Low priority) [5]. For each type of cancer, the initial treatment is given high priority while considering the degree of epidemic spread and the degree of shortage of human/material resources [5,6].

In Japan, the number of patients with COVID-19 has increased since the Japanese government declared a state of emergency without urban lockdown on April 7, 2020 [7]. The increase of COVID-19 patients has caused a temporary shortage of personal protective equipment (PPE) and an increase in hospital-acquired infections [8]. Therefore, it brings serious concerns about shortages of material/human resources, Japanese academic societies announced the recommendation of treatment for cancer in the COVID-19 pandemic, as have academic societies worldwide [9,10]. Previous studies have indicated that the COVID-19 pandemic has had an impact on cancer treatment, such as a decrease in the number of operations for patients with malignant tumors [11,12]. However, few studies have reported general effects, such as a decrease of hospital admissions related to the impact of the COVID-19 pandemic in the field of oncology. The aim of our study was to reveal the impact of the COVID-19 outbreak on the clinical practice of various cancers.

## METHODS

### Data source and study participants

We analyzed Diagnosis Procedure Combination (DPC) data from the Quality Indicator/Improvement Project (QIP) database, which was administered by the Department of Healthcare Economics and Quality Management, Kyoto University. The QIP database has DPC data from acute care hospitals voluntarily participating in the project. The number of participant hospitals was over 500, which took place all over Japan and included both public and private hospitals [13]. In Japan, the DPC/Per-Diem Payment System (PDPS) is a prospective payment system adopted for acute care hospitals. The number of general beds applied with the DPC/PDPS in 2018 was 482,618, which was 54% of all general beds of Japanese hospitals (482,618/891,872) [14,15]. The DPC data contain claims and clinical summary data, which include hospital identifiers, admission and discharge statuses (hospital death or not), cause of admission, main diagnosis, the most and second-most medical resource-intensive diagnoses, up to 10 comorbidities, and 10 complications. All diagnoses were classified based on the International Classification of Diseases, 10^th^ Revision (ICD-10) codes. For further details, refer to the following published paper [13].

We included patients who were aged 18 years or older and admitted between July 1, 2018 and June 30, 2020 and whose both major and most most-medical resource-intensive diagnosis was a cancer diagnosis based on ICD-10 codes. Among these cases, we analyzed in detail the top 12 most common types of cancer in Japan (colon/rectum, lung, gastric, breast, bladder & urinary tract, pancreas, non-Hodgkin lymphoma, liver, prostate, esophagus, uterus, and gallbladder & biliary tract) based on the WHO’s research [16]. We used claims codes for surgical procedures classification codes from the Ministry of Health and Welfare in Japan for identifying the surgical procedure.

### Statistical analyses

We divided the population into two: those who were admitted and discharged between July 2018 and June 2019 and between July 2019 and June 2020 for year-over-year comparisons. Because the DPC data were coded on the day of discharge, the data of patients who were not discharged before the end of June 2020 were not available, even though their date of admission was before June 30, 2020. We then described the total number of admission cases, the cases with any radical surgical procedure, and cases with chemotherapy for each cancer. Furthermore, we assessed the monthly ratio of the proportion of admission cases with each cancer diagnosis to the total number of cancer cases.

The ratio of proportions = proportion of admission cases [2019/07-2020/06] / proportion of admission cases [2018/07-2019/06]

where

proportion of admission cases [2019/7-2020/6] = number of admission cases with each diagnosis of cancer per month between July 2019 and June 2020 / number of admission cases with all cancers per month between July 2019 and June 2020

and where

proportion of admission cases [2018/7-2019/6] = number of admission cases with each diagnosis of cancer per month between July 2018 and June 2019 / number of admission cases with all cancers per month between July 2018 and June 2019.

Continuous variables were presented as median, interquartile range, and categorical variables were presented as numbers and proportions. We performed an interrupted time series (ITS) analysis, with segmented and Poisson regressions, taking into account seasonality, secular trends, and overdispersion of data, to evaluate the impact of the COVID-19 pandemic on population-level admissions [17,18]. Seasonality was considered by including harmonic terms (sines and cosines) with 12-month periods to our model [17]. The validity of the Poisson regression model was assessed by checking the correlograms (functions of autocorrelation and partial autocorrelation) and the residuals.

The Japanese government declared a state of emergency on April 7, 2020, and because our study examined the number of admission cases per month, we designated April 2020 as the beginning of intervention and estimated that the declaration of emergency with COVID-19 pandemic would rapidly decrease the admission volume after April 7, 2020 [7]. Because the DPC data were generated on the day of discharge, our model assessed the change of monthly admission volume based on the date of discharge.

Statistical significance was regarded as a two-sided P-value <0.05. All analyses were performed using R 3.6.3 (R Foundation for Statistical Computing, Vienna, Austria).

### Ethical considerations

The present study was approved by the ethics committee of Kyoto University (approval number: R0135), and we did not require informed consent because of the use of anonymized data, in accordance with the Ethical Guidelines for Medical and Health Research Involving Human Subjects, as stipulated by the Japanese Government. We declare that there are no conflicts of interest in the manuscript.

## RESULTS

We included a total of 403,344 cases with any cancer diagnosis, in accordance with the top 12 most common types of cancer during the study period. The median age was 72 years old, and 59% of the study population were males. The most common primary lesion was colorectal (20.5%), followed by lung (17.5%) and stomach (12.3%). Patients who underwent any radical surgery or any chemotherapy were 33.7% and 36.4%, respectively (Table1). The characteristics of the study population with each cancer diagnosis case were described in Supplementary Table1.

**Table1.** Baseline characteristics of study population

| Characteristic | Overall (n = 403,344) |  |
| --- | --- | --- |
| **Age** |  |  |
| Median (IQR) | 72 | (65, 78) |
| **Age category** |  |  |
| 75 years ≥ | 161,499 | 40.0% |
| 65 – 74 years | 145,631 | 36.1% |
| 64 years ≤ | 96,214 | 23.9% |
| **Sex** |  |  |
| Males | 239,345 | 59.3% |
| Females | 163,999 | 40.7% |
| **Primary legion** |  |  |
| Colon/rectum | 82,541 | 20.5% |
| Lung | 70,455 | 17.5% |
| Stomach | 49,504 | 12.3% |
| Breast | 32,662 | 8.1% |
| Bladder & Urinary tract | 32,186 | 8.0% |
| Pancreas | 25,948 | 6.4% |
| Non-Hodgkin lymphoma | 25,273 | 6.3% |
| Liver | 25,588 | 6.3% |
| Prostate | 16,045 | 4.0% |
| Esophagus | 15,668 | 3.9% |
| Uterus | 14,618 | 3.6% |
| Gallbladder & Biliary tract | 12,856 | 3.2% |
| **Primary/Recurrent** |  |  |
| Primary | 290,968 | 72.1% |
| Recurrent | 101,501 | 25.2% |
| Unknown | 10,875 | 2.7% |
| **Purpose of admission** |  |  |
| Special purpose | 392,105 | 97.2% |
| Examination | 11,239 | 2.8% |
| **Anti-cancer treatment** |  |  |
| Any radical surgery | 135,815 | 33.7% |
| Any chemotherapy | 146,977 | 36.4% |
| Length of hospital stay, median (IQR) | 9.0 | (4.0, 16.0) |
| In-hospital mortality | 35,015 | 8.7% |

Abbreviations: IQR: Interquartile Range

For almost all cancer cases, the number of admission cases decreased in May 2020 compared with May 2019. In particular, colorectal, lung, gastric, breast, uterine, and esophageal cancer cases decreased by over 10% (Table2). The monthly ratio of the proportion of each cancer case to all cancer cases was near 1. However, only the proportion of gastric cancer cases decreased (Table2). The trends in the number of cases for each cancer diagnosis are shown in Supplementary Figure1. The number of cases in June largely decreased because the study population contained the cases that were admitted and discharged by the end of June.

**Table2.**
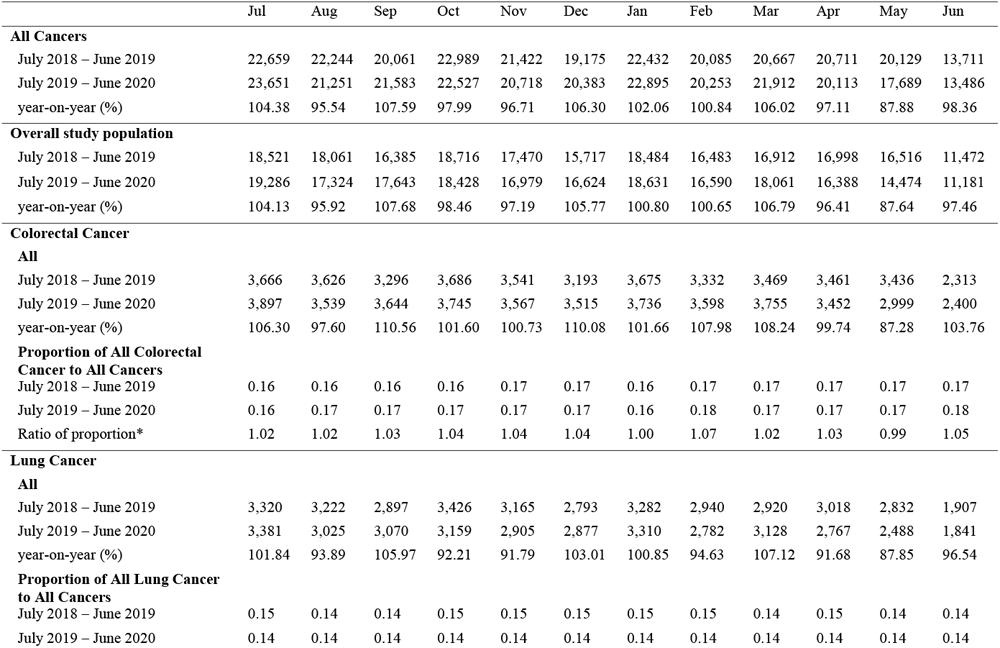
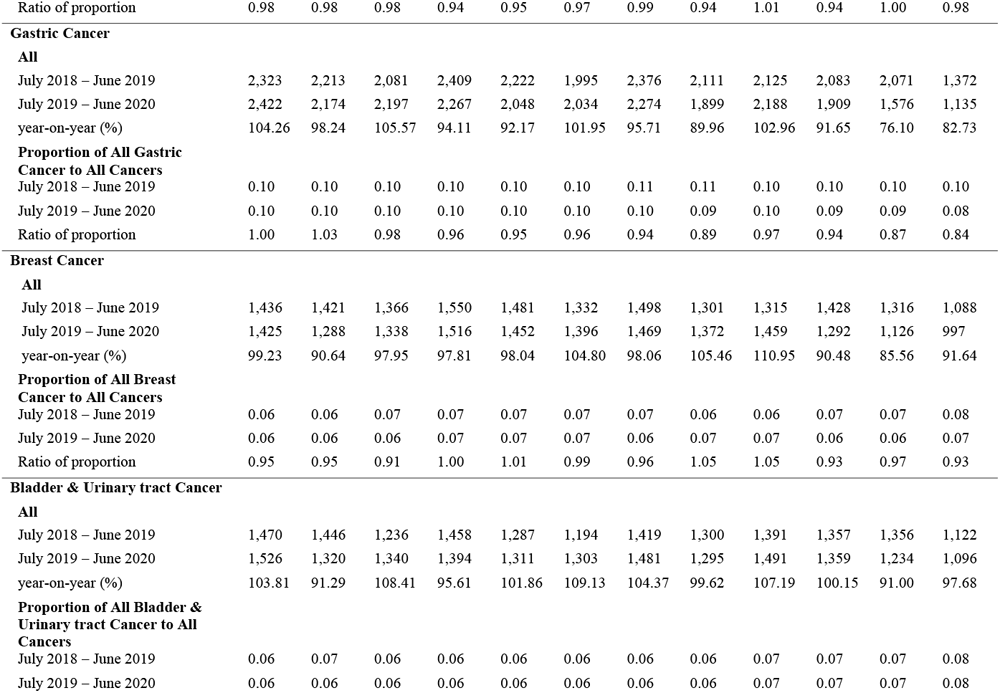
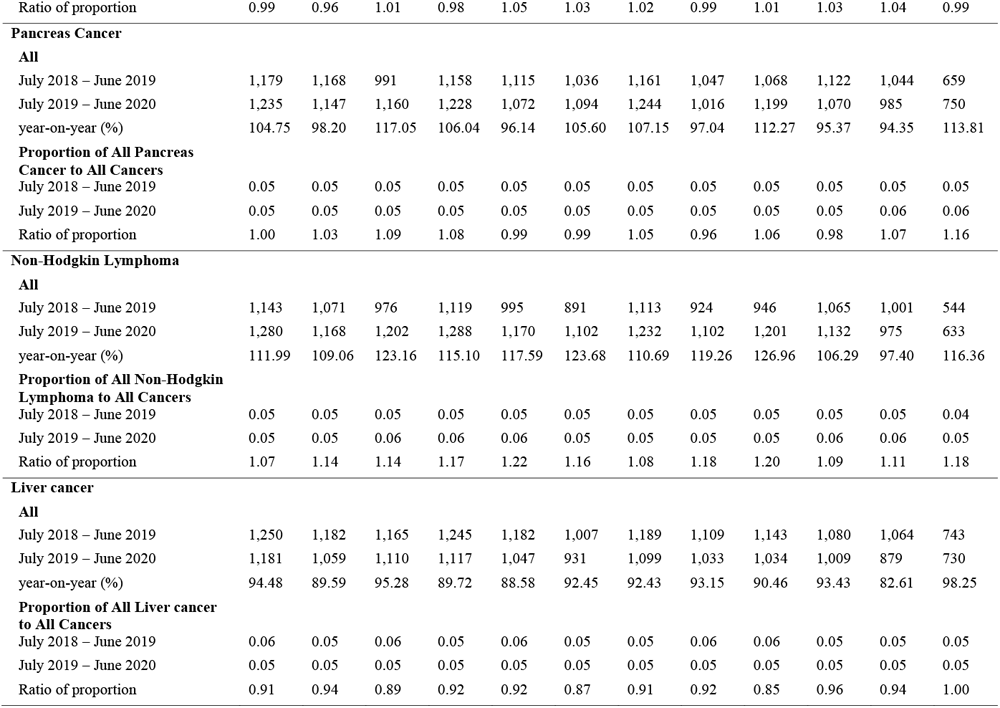
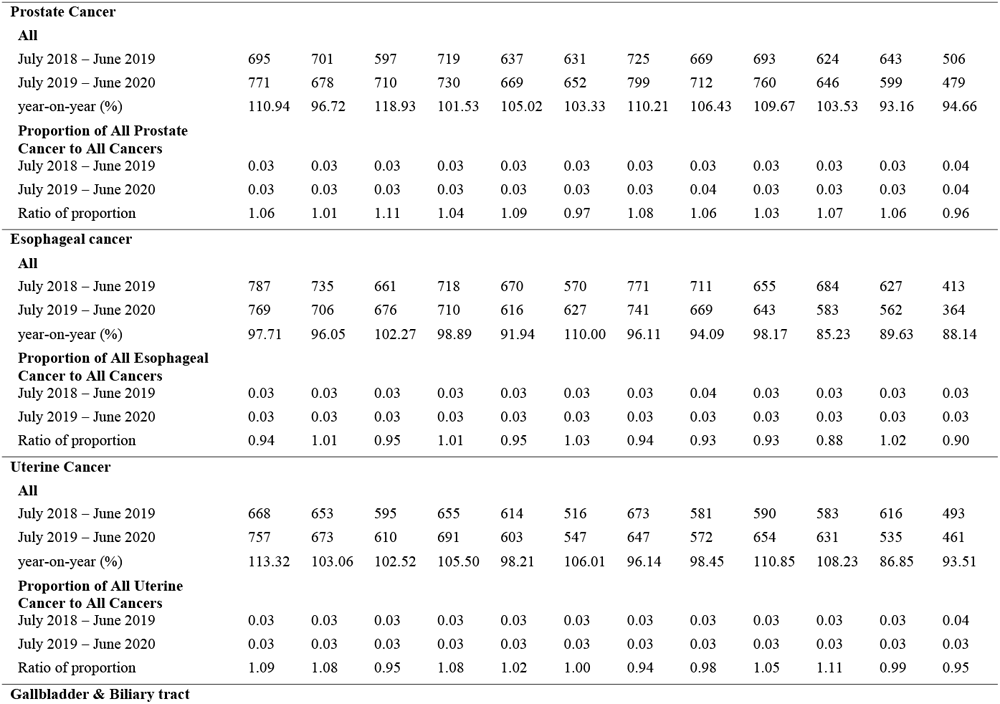
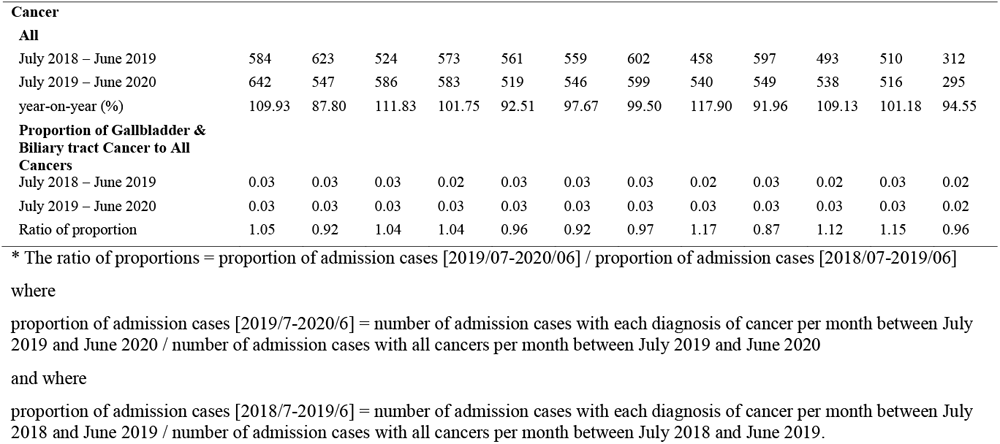
Number of admission cases per month for each cancer diagnosis

The range of decrease of cases with any radical surgical procedure and cases with chemotherapy varied depending on the type of cancer (Table3). In May 2020, the number of cases with colorectal, lung, gastric, or esophageal cancer with surgery decreased by more than 10% compared with those in the previous year. The number of cases for chemotherapy with lung, gastric, breast, or uterus cancer decreased by more than 10% compared with those in the previous year. In contrast, cases of non-Hodgkin lymphoma, prostate, pancreas, biliary duct cancer reduction in hospital admissions was less than 10% in May 2020 compared with other types of cancer.

**Table3.**
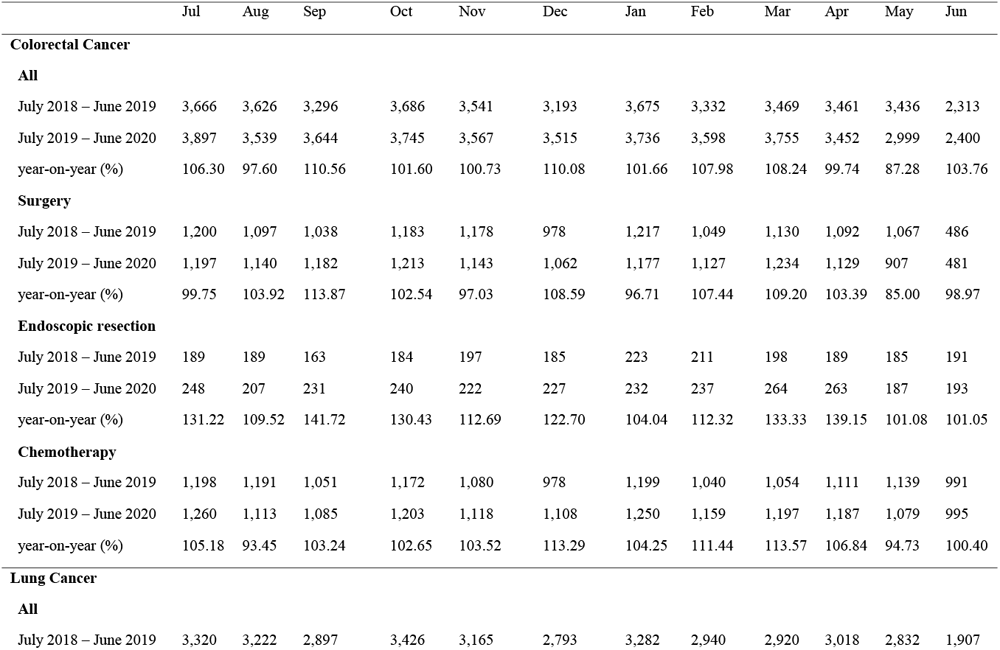
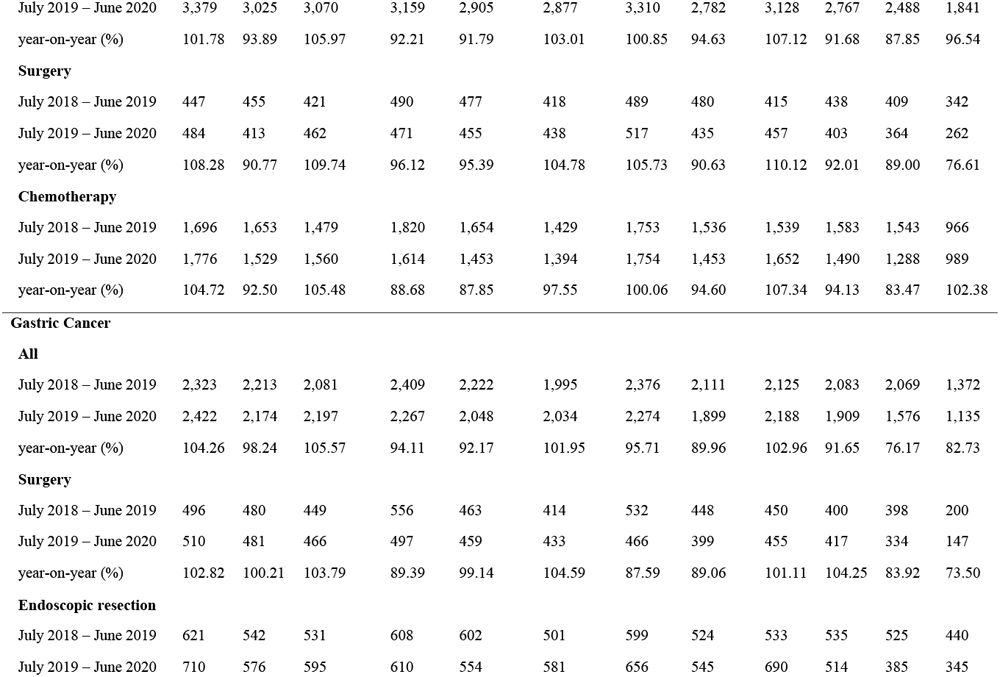
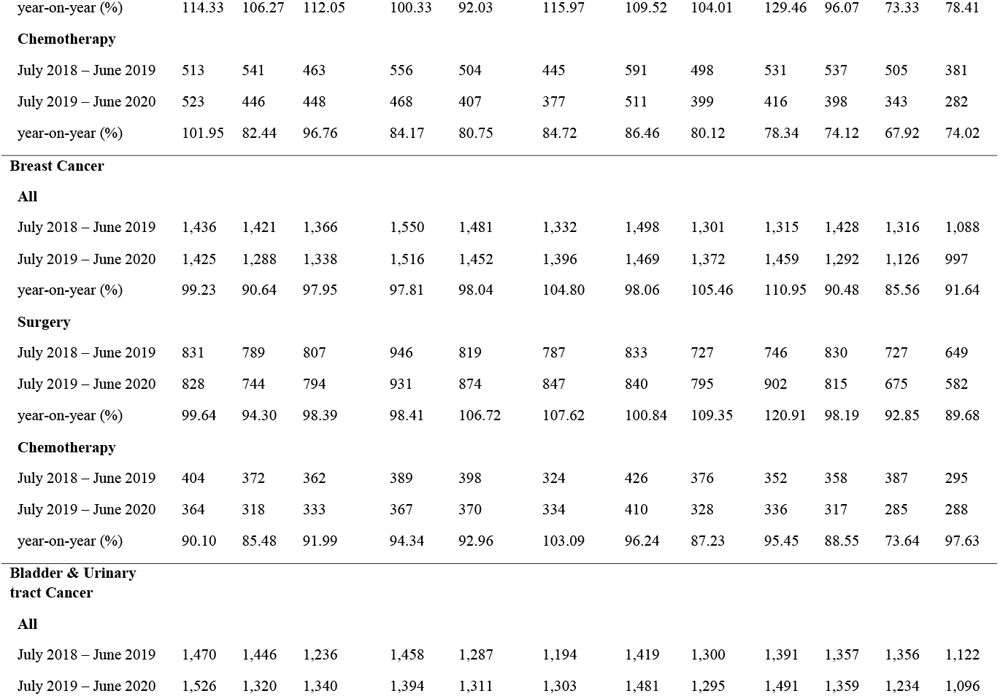
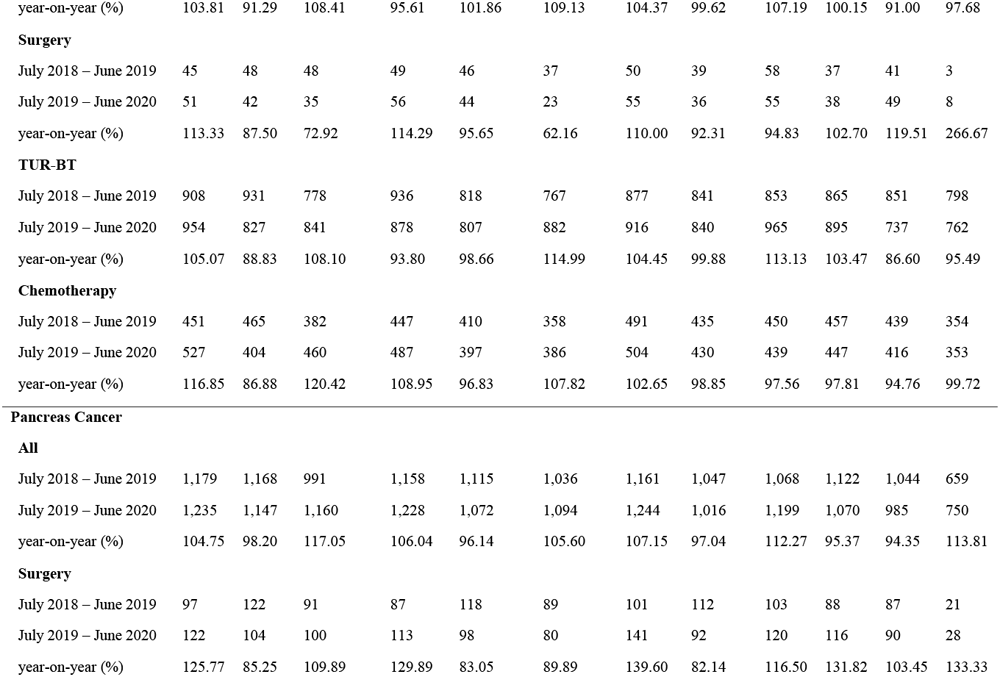
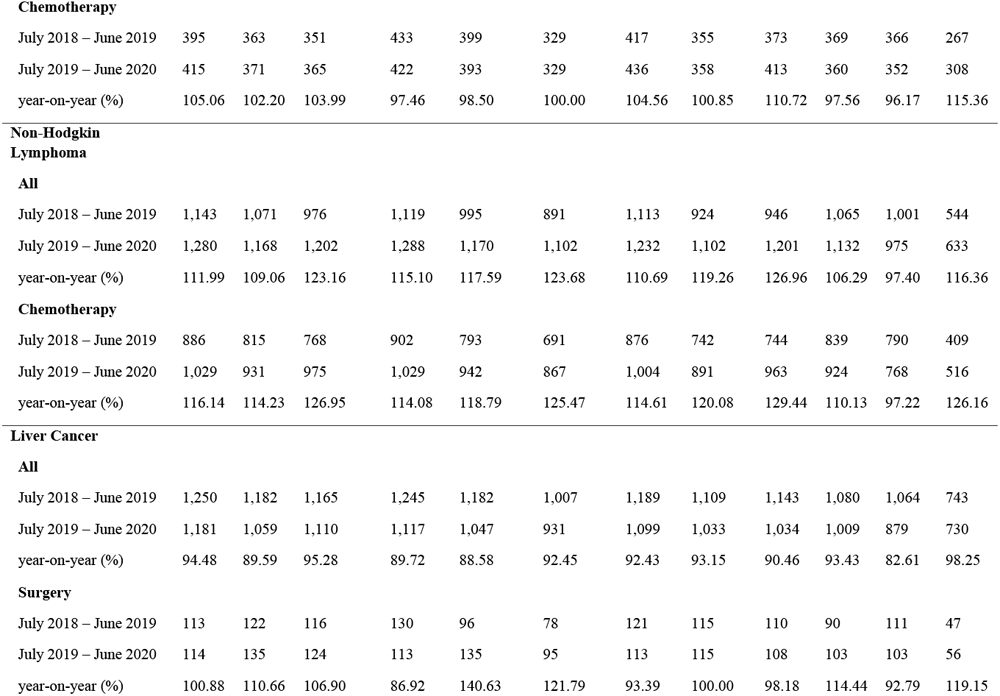
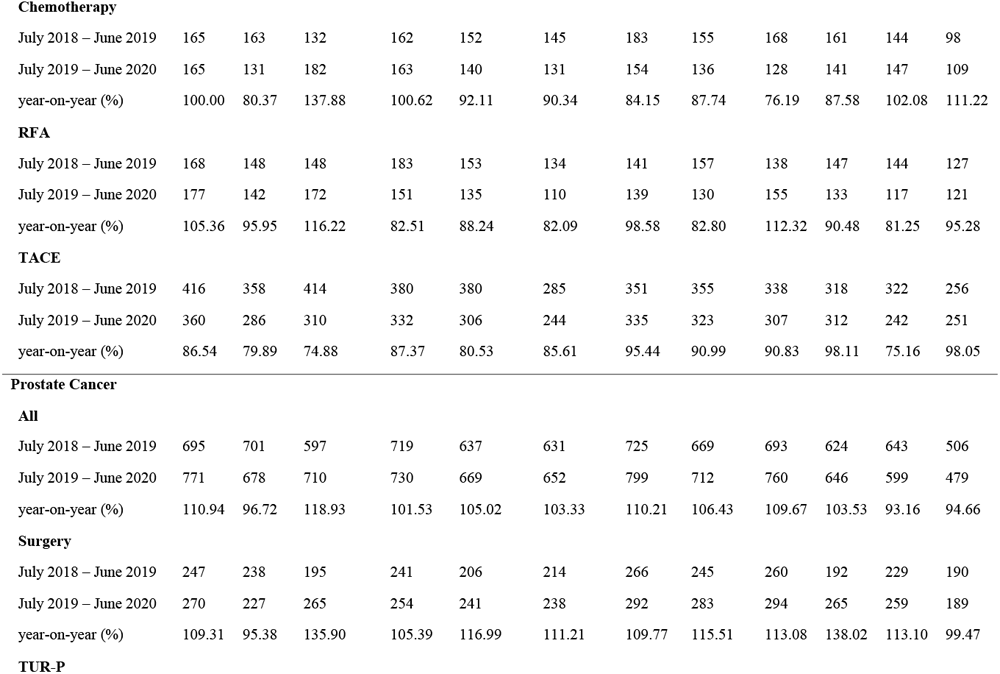
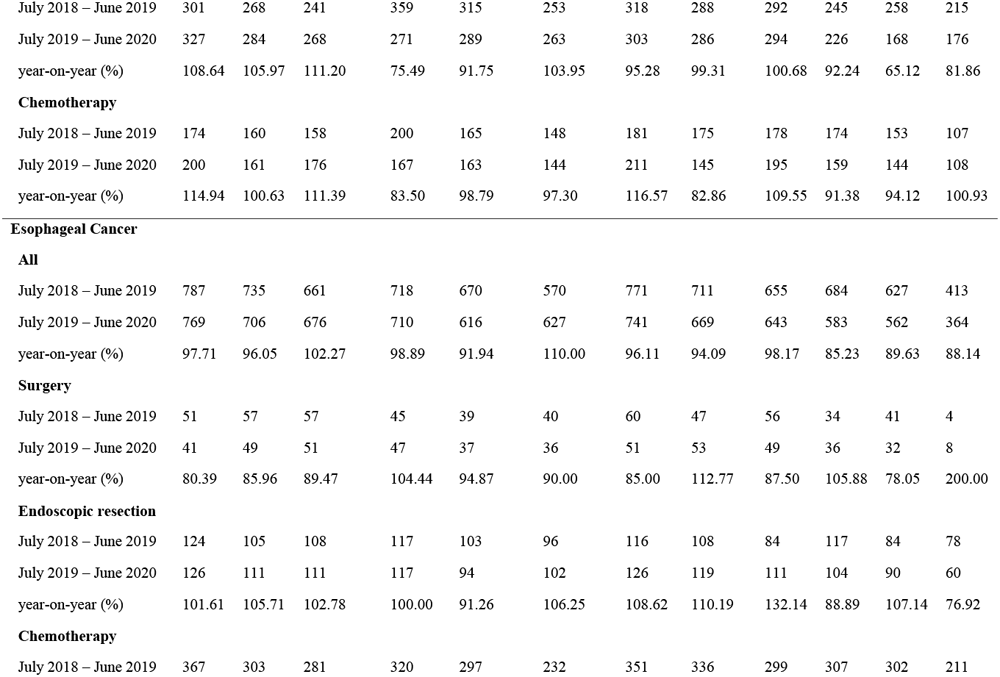
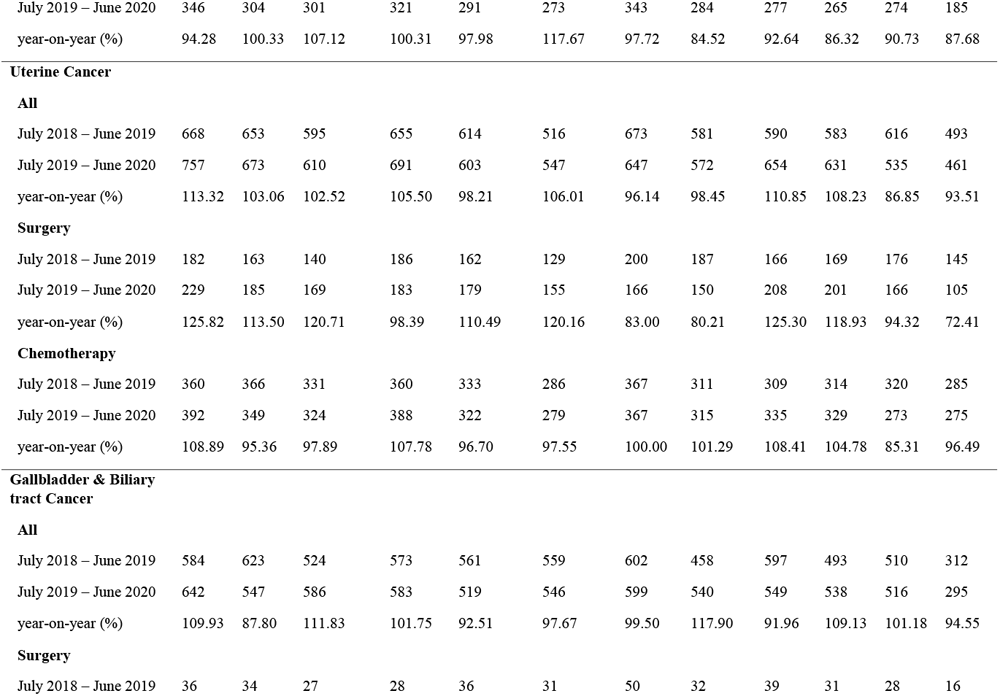
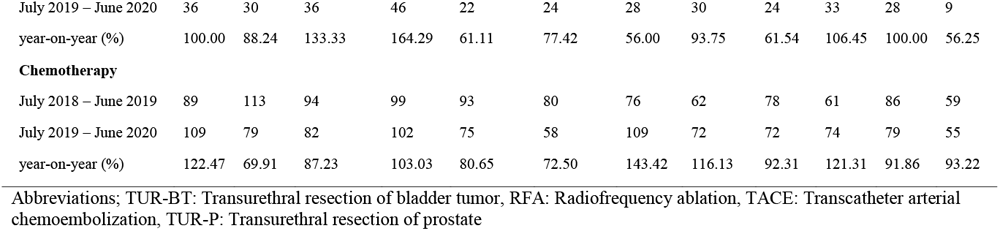
Number of monthly admission cases for each treatment for each cancer diagnosis

|  | Jul | Aug | Sep | Oct | Nov | Dec | Jan | Feb | Mar | Apr | May | Jun |
| --- | --- | --- | --- | --- | --- | --- | --- | --- | --- | --- | --- | --- |
| **Colorectal Cancer** |  |  |  |  |  |  |  |  |  |  |  |  |
| **All** |  |  |  |  |  |  |  |  |  |  |  |  |
| July 2018 – June 2019 | 3,666 | 3,626 | 3,296 | 3,686 | 3,541 | 3,193 | 3,675 | 3,332 | 3,469 | 3,461 | 3,436 | 2,313 |
| July 2019 – June 2020 | 3,897 | 3,539 | 3,644 | 3,745 | 3,567 | 3,515 | 3,736 | 3,598 | 3,755 | 3,452 | 2,999 | 2,400 |
| year-on-year (%) | 106.30 | 97.60 | 110.56 | 101.60 | 100.73 | 110.08 | 101.66 | 107.98 | 108.24 | 99.74 | 87.28 | 103.76 |
| **Surgery** |  |  |  |  |  |  |  |  |  |  |  |  |
| July 2018 – June 2019 | 1,200 | 1,097 | 1,038 | 1,183 | 1,178 | 978 | 1,217 | 1,049 | 1,130 | 1,092 | 1,067 | 486 |
| July 2019 – June 2020 | 1,197 | 1,140 | 1,182 | 1,213 | 1,143 | 1,062 | 1,177 | 1,127 | 1,234 | 1,129 | 907 | 481 |
| year-on-year (%) | 99.75 | 103.92 | 113.87 | 102.54 | 97.03 | 108.59 | 96.71 | 107.44 | 109.20 | 103.39 | 85.00 | 98.97 |
| **Endoscopic resection** |  |  |  |  |  |  |  |  |  |  |  |  |
| July 2018 – June 2019 | 189 | 189 | 163 | 184 | 197 | 185 | 223 | 211 | 198 | 189 | 185 | 191 |
| July 2019 – June 2020 | 248 | 207 | 231 | 240 | 222 | 227 | 232 | 237 | 264 | 263 | 187 | 193 |
| year-on-year (%) | 131.22 | 109.52 | 141.72 | 130.43 | 112.69 | 122.70 | 104.04 | 112.32 | 133.33 | 139.15 | 101.08 | 101.05 |
| **Chemotherapy** |  |  |  |  |  |  |  |  |  |  |  |  |
| July 2018 – June 2019 | 1,198 | 1,191 | 1,051 | 1,172 | 1,080 | 978 | 1,199 | 1,040 | 1,054 | 1,111 | 1,139 | 991 |
| July 2019 – June 2020 | 1,260 | 1,113 | 1,085 | 1,203 | 1,118 | 1,108 | 1,250 | 1,159 | 1,197 | 1,187 | 1,079 | 995 |
| year-on-year (%) | 105.18 | 93.45 | 103.24 | 102.65 | 103.52 | 113.29 | 104.25 | 111.44 | 113.57 | 106.84 | 94.73 | 100.40 |
| **Lung Cancer** |  |  |  |  |  |  |  |  |  |  |  |  |
| **All** |  |  |  |  |  |  |  |  |  |  |  |  |
| July 2018 – June 2019 | 3,320 | 3,222 | 2,897 | 3,426 | 3,165 | 2,793 | 3,282 | 2,940 | 2,920 | 3,018 | 2,832 | 1,907 |
| July 2019 – June 2020 | 3,379 | 3,025 | 3,070 | 3,159 | 2,905 | 2,877 | 3,310 | 2,782 | 3,128 | 2,767 | 2,488 | 1,841 |
| year-on-year (%) | 101.78 | 93.89 | 105.97 | 92.21 | 91.79 | 103.01 | 100.85 | 94.63 | 107.12 | 91.68 | 87.85 | 96.54 |
| **Surgery** |  |  |  |  |  |  |  |  |  |  |  |  |
| July 2018 – June 2019 | 447 | 455 | 421 | 490 | 477 | 418 | 489 | 480 | 415 | 438 | 409 | 342 |
| July 2019 – June 2020 | 484 | 413 | 462 | 471 | 455 | 438 | 517 | 435 | 457 | 403 | 364 | 262 |
| year-on-year (%) | 108.28 | 90.77 | 109.74 | 96.12 | 95.39 | 104.78 | 105.73 | 90.63 | 110.12 | 92.01 | 89.00 | 76.61 |
| **Chemotherapy** |  |  |  |  |  |  |  |  |  |  |  |  |
| July 2018 – June 2019 | 1,696 | 1,653 | 1,479 | 1,820 | 1,654 | 1,429 | 1,753 | 1,536 | 1,539 | 1,583 | 1,543 | 966 |
| July 2019 – June 2020 | 1,776 | 1,529 | 1,560 | 1,614 | 1,453 | 1,394 | 1,754 | 1,453 | 1,652 | 1,490 | 1,288 | 989 |
| year-on-year (%) | 104.72 | 92.50 | 105.48 | 88.68 | 87.85 | 97.55 | 100.06 | 94.60 | 107.34 | 94.13 | 83.47 | 102.38 |
| **Gastric Cancer** |  |  |  |  |  |  |  |  |  |  |  |  |
| **All** |  |  |  |  |  |  |  |  |  |  |  |  |
| July 2018 – June 2019 | 2,323 | 2,213 | 2,081 | 2,409 | 2,222 | 1,995 | 2,376 | 2,111 | 2,125 | 2,083 | 2,069 | 1,372 |
| July 2019 – June 2020 | 2,422 | 2,174 | 2,197 | 2,267 | 2,048 | 2,034 | 2,274 | 1,899 | 2,188 | 1,909 | 1,576 | 1,135 |
| year-on-year (%) | 104.26 | 98.24 | 105.57 | 94.11 | 92.17 | 101.95 | 95.71 | 89.96 | 102.96 | 91.65 | 76.17 | 82.73 |
| **Surgery** |  |  |  |  |  |  |  |  |  |  |  |  |
| July 2018 – June 2019 | 496 | 480 | 449 | 556 | 463 | 414 | 532 | 448 | 450 | 400 | 398 | 200 |
| July 2019 – June 2020 | 510 | 481 | 466 | 497 | 459 | 433 | 466 | 399 | 455 | 417 | 334 | 147 |
| year-on-year (%) | 102.82 | 100.21 | 103.79 | 89.39 | 99.14 | 104.59 | 87.59 | 89.06 | 101.11 | 104.25 | 83.92 | 73.50 |
| **Endoscopic resection** |  |  |  |  |  |  |  |  |  |  |  |  |
| July 2018 – June 2019 | 621 | 542 | 531 | 608 | 602 | 501 | 599 | 524 | 533 | 535 | 525 | 440 |
| July 2019 – June 2020 | 710 | 576 | 595 | 610 | 554 | 581 | 656 | 545 | 690 | 514 | 385 | 345 |
| year-on-year (%) | 114.33 | 106.27 | 112.05 | 100.33 | 92.03 | 115.97 | 109.52 | 104.01 | 129.46 | 96.07 | 73.33 | 78.41 |
| **Chemotherapy** |  |  |  |  |  |  |  |  |  |  |  |  |
| July 2018 – June 2019 | 513 | 541 | 463 | 556 | 504 | 445 | 591 | 498 | 531 | 537 | 505 | 381 |
| July 2019 – June 2020 | 523 | 446 | 448 | 468 | 407 | 377 | 511 | 399 | 416 | 398 | 343 | 282 |
| year-on-year (%) | 101.95 | 82.44 | 96.76 | 84.17 | 80.75 | 84.72 | 86.46 | 80.12 | 78.34 | 74.12 | 67.92 | 74.02 |
| **Breast Cancer** |  |  |  |  |  |  |  |  |  |  |  |  |
| **All** |  |  |  |  |  |  |  |  |  |  |  |  |
| July 2018 – June 2019 | 1,436 | 1,421 | 1,366 | 1,550 | 1,481 | 1,332 | 1,498 | 1,301 | 1,315 | 1,428 | 1,316 | 1,088 |
| July 2019 – June 2020 | 1,425 | 1,288 | 1,338 | 1,516 | 1,452 | 1,396 | 1,469 | 1,372 | 1,459 | 1,292 | 1,126 | 997 |
| year-on-year (%) | 99.23 | 90.64 | 97.95 | 97.81 | 98.04 | 104.80 | 98.06 | 105.46 | 110.95 | 90.48 | 85.56 | 91.64 |
| **Surgery** |  |  |  |  |  |  |  |  |  |  |  |  |
| July 2018 – June 2019 | 831 | 789 | 807 | 946 | 819 | 787 | 833 | 727 | 746 | 830 | 727 | 649 |
| July 2019 – June 2020 | 828 | 744 | 794 | 931 | 874 | 847 | 840 | 795 | 902 | 815 | 675 | 582 |
| year-on-year (%) | 99.64 | 94.30 | 98.39 | 98.41 | 106.72 | 107.62 | 100.84 | 109.35 | 120.91 | 98.19 | 92.85 | 89.68 |
| **Chemotherapy** |  |  |  |  |  |  |  |  |  |  |  |  |
| July 2018 – June 2019 | 404 | 372 | 362 | 389 | 398 | 324 | 426 | 376 | 352 | 358 | 387 | 295 |
| July 2019 – June 2020 | 364 | 318 | 333 | 367 | 370 | 334 | 410 | 328 | 336 | 317 | 285 | 288 |
| year-on-year (%) | 90.10 | 85.48 | 91.99 | 94.34 | 92.96 | 103.09 | 96.24 | 87.23 | 95.45 | 88.55 | 73.64 | 97.63 |
| **Bladder & Urinary tract Cancer** |  |  |  |  |  |  |  |  |  |  |  |  |
| **All** |  |  |  |  |  |  |  |  |  |  |  |  |
| July 2018 – June 2019 | 1,470 | 1,446 | 1,236 | 1,458 | 1,287 | 1,194 | 1,419 | 1,300 | 1,391 | 1,357 | 1,356 | 1,122 |
| July 2019 – June 2020 | 1,526 | 1,320 | 1,340 | 1,394 | 1,311 | 1,303 | 1,481 | 1,295 | 1,491 | 1,359 | 1,234 | 1,096 |
| year-on-year (%) | 103.81 | 91.29 | 108.41 | 95.61 | 101.86 | 109.13 | 104.37 | 99.62 | 107.19 | 100.15 | 91.00 | 97.68 |
| **Surgery** |  |  |  |  |  |  |  |  |  |  |  |  |
| July 2018 – June 2019 | 45 | 48 | 48 | 49 | 46 | 37 | 50 | 39 | 58 | 37 | 41 | 3 |
| July 2019 – June 2020 | 51 | 42 | 35 | 56 | 44 | 23 | 55 | 36 | 55 | 38 | 49 | 8 |
| year-on-year (%) | 113.33 | 87.50 | 72.92 | 114.29 | 95.65 | 62.16 | 110.00 | 92.31 | 94.83 | 102.70 | 119.51 | 266.67 |
| **TUR-BT** |  |  |  |  |  |  |  |  |  |  |  |  |
| July 2018 – June 2019 | 908 | 931 | 778 | 936 | 818 | 767 | 877 | 841 | 853 | 865 | 851 | 798 |
| July 2019 – June 2020 | 954 | 827 | 841 | 878 | 807 | 882 | 916 | 840 | 965 | 895 | 737 | 762 |
| year-on-year (%) | 105.07 | 88.83 | 108.10 | 93.80 | 98.66 | 114.99 | 104.45 | 99.88 | 113.13 | 103.47 | 86.60 | 95.49 |
| **Chemotherapy** |  |  |  |  |  |  |  |  |  |  |  |  |
| July 2018 – June 2019 | 451 | 465 | 382 | 447 | 410 | 358 | 491 | 435 | 450 | 457 | 439 | 354 |
| July 2019 – June 2020 | 527 | 404 | 460 | 487 | 397 | 386 | 504 | 430 | 439 | 447 | 416 | 353 |
| year-on-year (%) | 116.85 | 86.88 | 120.42 | 108.95 | 96.83 | 107.82 | 102.65 | 98.85 | 97.56 | 97.81 | 94.76 | 99.72 |
| **Pancreas Cancer** |  |  |  |  |  |  |  |  |  |  |  |  |
| **All** |  |  |  |  |  |  |  |  |  |  |  |  |
| July 2018 – June 2019 | 1,179 | 1,168 | 991 | 1,158 | 1,115 | 1,036 | 1,161 | 1,047 | 1,068 | 1,122 | 1,044 | 659 |
| July 2019 – June 2020 | 1,235 | 1,147 | 1,160 | 1,228 | 1,072 | 1,094 | 1,244 | 1,016 | 1,199 | 1,070 | 985 | 750 |
| year-on-year (%) | 104.75 | 98.20 | 117.05 | 106.04 | 96.14 | 105.60 | 107.15 | 97.04 | 112.27 | 95.37 | 94.35 | 113.81 |
| **Surgery** |  |  |  |  |  |  |  |  |  |  |  |  |
| July 2018 – June 2019 | 97 | 122 | 91 | 87 | 118 | 89 | 101 | 112 | 103 | 88 | 87 | 21 |
| July 2019 – June 2020 | 122 | 104 | 100 | 113 | 98 | 80 | 141 | 92 | 120 | 116 | 90 | 28 |
| year-on-year (%) | 125.77 | 85.25 | 109.89 | 129.89 | 83.05 | 89.89 | 139.60 | 82.14 | 116.50 | 131.82 | 103.45 | 133.33 |
| **Chemotherapy** |  |  |  |  |  |  |  |  |  |  |  |  |
| July 2018 – June 2019 | 395 | 363 | 351 | 433 | 399 | 329 | 417 | 355 | 373 | 369 | 366 | 267 |
| July 2019 – June 2020 | 415 | 371 | 365 | 422 | 393 | 329 | 436 | 358 | 413 | 360 | 352 | 308 |
| year-on-year (%) | 105.06 | 102.20 | 103.99 | 97.46 | 98.50 | 100.00 | 104.56 | 100.85 | 110.72 | 97.56 | 96.17 | 115.36 |
| July 2018 – June 2019 | 1,143 | 1,071 | 976 | 1,119 | 995 | 891 | 1,113 | 924 | 946 | 1,065 | 1,001 | 544 |
| July 2019 – June 2020 | 1,280 | 1,168 | 1,202 | 1,288 | 1,170 | 1,102 | 1,232 | 1,102 | 1,201 | 1,132 | 975 | 633 |
| year-on-year (%) | 111.99 | 109.06 | 123.16 | 115.10 | 117.59 | 123.68 | 110.69 | 119.26 | 126.96 | 106.29 | 97.40 | 116.36 |
| **Chemotherapy** |  |  |  |  |  |  |  |  |  |  |  |  |
| July 2018 – June 2019 | 886 | 815 | 768 | 902 | 793 | 691 | 876 | 742 | 744 | 839 | 790 | 409 |
| July 2019 – June 2020 | 1,029 | 931 | 975 | 1,029 | 942 | 867 | 1,004 | 891 | 963 | 924 | 768 | 516 |
| year-on-year (%) | 116.14 | 114.23 | 126.95 | 114.08 | 118.79 | 125.47 | 114.61 | 120.08 | 129.44 | 110.13 | 97.22 | 126.16 |
| **Liver Cancer****All** |  |  |  |  |  |  |  |  |  |  |  |  |
| July 2018 – June 2019 | 1,250 | 1,182 | 1,165 | 1,245 | 1,182 | 1,007 | 1,189 | 1,109 | 1,143 | 1,080 | 1,064 | 743 |
| July 2019 – June 2020 | 1,181 | 1,059 | 1,110 | 1,117 | 1,047 | 931 | 1,099 | 1,033 | 1,034 | 1,009 | 879 | 730 |
| year-on-year (%) | 94.48 | 89.59 | 95.28 | 89.72 | 88.58 | 92.45 | 92.43 | 93.15 | 90.46 | 93.43 | 82.61 | 98.25 |
| **Surgery** |  |  |  |  |  |  |  |  |  |  |  |  |
| July 2018 – June 2019 | 113 | 122 | 116 | 130 | 96 | 78 | 121 | 115 | 110 | 90 | 111 | 47 |
| July 2019 – June 2020 | 114 | 135 | 124 | 113 | 135 | 95 | 113 | 115 | 108 | 103 | 103 | 56 |
| year-on-year (%) | 100.88 | 110.66 | 106.90 | 86.92 | 140.63 | 121.79 | 93.39 | 100.00 | 98.18 | 114.44 | 92.79 | 119.15 |
| **Chemotherapy** |  |  |  |  |  |  |  |  |  |  |  |  |
| July 2018 – June 2019 | 165 | 163 | 132 | 162 | 152 | 145 | 183 | 155 | 168 | 161 | 144 | 98 |
| July 2019 – June 2020 | 165 | 131 | 182 | 163 | 140 | 131 | 154 | 136 | 128 | 141 | 147 | 109 |
| year-on-year (%) | 100.00 | 80.37 | 137.88 | 100.62 | 92.11 | 90.34 | 84.15 | 87.74 | 76.19 | 87.58 | 102.08 | 111.22 |
| **RFA** |  |  |  |  |  |  |  |  |  |  |  |  |
| July 2018 – June 2019 | 168 | 148 | 148 | 183 | 153 | 134 | 141 | 157 | 138 | 147 | 144 | 127 |
| July 2019 – June 2020 | 177 | 142 | 172 | 151 | 135 | 110 | 139 | 130 | 155 | 133 | 117 | 121 |
| year-on-year (%) | 105.36 | 95.95 | 116.22 | 82.51 | 88.24 | 82.09 | 98.58 | 82.80 | 112.32 | 90.48 | 81.25 | 95.28 |
| **TACE** |  |  |  |  |  |  |  |  |  |  |  |  |
| July 2018 – June 2019 | 416 | 358 | 414 | 380 | 380 | 285 | 351 | 355 | 338 | 318 | 322 | 256 |
| July 2019 – June 2020 | 360 | 286 | 310 | 332 | 306 | 244 | 335 | 323 | 307 | 312 | 242 | 251 |
| year-on-year (%) | 86.54 | 79.89 | 74.88 | 87.37 | 80.53 | 85.61 | 95.44 | 90.99 | 90.83 | 98.11 | 75.16 | 98.05 |
| **Prostate Cancer****All** |  |  |  |  |  |  |  |  |  |  |  |  |
| July 2018 – June 2019 | 695 | 701 | 597 | 719 | 637 | 631 | 725 | 669 | 693 | 624 | 643 | 506 |
| July 2019 – June 2020 | 771 | 678 | 710 | 730 | 669 | 652 | 799 | 712 | 760 | 646 | 599 | 479 |
| year-on-year (%) | 110.94 | 96.72 | 118.93 | 101.53 | 105.02 | 103.33 | 110.21 | 106.43 | 109.67 | 103.53 | 93.16 | 94.66 |
| **Surgery** |  |  |  |  |  |  |  |  |  |  |  |  |
| July 2018 – June 2019 | 247 | 238 | 195 | 241 | 206 | 214 | 266 | 245 | 260 | 192 | 229 | 190 |
| July 2019 – June 2020 | 270 | 227 | 265 | 254 | 241 | 238 | 292 | 283 | 294 | 265 | 259 | 189 |
| year-on-year (%) | 109.31 | 95.38 | 135.90 | 105.39 | 116.99 | 111.21 | 109.77 | 115.51 | 113.08 | 138.02 | 113.10 | 99.47 |
| **TUR-P** |  |  |  |  |  |  |  |  |  |  |  |  |
| July 2018 – June 2019 | 301 | 268 | 241 | 359 | 315 | 253 | 318 | 288 | 292 | 245 | 258 | 215 |
| July 2019 – June 2020 | 327 | 284 | 268 | 271 | 289 | 263 | 303 | 286 | 294 | 226 | 168 | 176 |
| year-on-year (%) | 108.64 | 105.97 | 111.20 | 75.49 | 91.75 | 103.95 | 95.28 | 99.31 | 100.68 | 92.24 | 65.12 | 81.86 |
| **Chemotherapy** |  |  |  |  |  |  |  |  |  |  |  |  |
| July 2018 – June 2019 | 174 | 160 | 158 | 200 | 165 | 148 | 181 | 175 | 178 | 174 | 153 | 107 |
| July 2019 – June 2020 | 200 | 161 | 176 | 167 | 163 | 144 | 211 | 145 | 195 | 159 | 144 | 108 |
| year-on-year (%) | 114.94 | 100.63 | 111.39 | 83.50 | 98.79 | 97.30 | 116.57 | 82.86 | 109.55 | 91.38 | 94.12 | 100.93 |
| **Esophageal Cancer** |  |  |  |  |  |  |  |  |  |  |  |  |
| **All** |  |  |  |  |  |  |  |  |  |  |  |  |
| July 2018 – June 2019 | 787 | 735 | 661 | 718 | 670 | 570 | 771 | 711 | 655 | 684 | 627 | 413 |
| July 2019 – June 2020 | 769 | 706 | 676 | 710 | 616 | 627 | 741 | 669 | 643 | 583 | 562 | 364 |
| year-on-year (%) | 97.71 | 96.05 | 102.27 | 98.89 | 91.94 | 110.00 | 96.11 | 94.09 | 98.17 | 85.23 | 89.63 | 88.14 |
| **Surgery** |  |  |  |  |  |  |  |  |  |  |  |  |
| July 2018 – June 2019 | 51 | 57 | 57 | 45 | 39 | 40 | 60 | 47 | 56 | 34 | 41 | 4 |
| July 2019 – June 2020 | 41 | 49 | 51 | 47 | 37 | 36 | 51 | 53 | 49 | 36 | 32 | 8 |
| year-on-year (%) | 80.39 | 85.96 | 89.47 | 104.44 | 94.87 | 90.00 | 85.00 | 112.77 | 87.50 | 105.88 | 78.05 | 200.00 |
| **Endoscopic resection** |  |  |  |  |  |  |  |  |  |  |  |  |
| July 2018 – June 2019 | 124 | 105 | 108 | 117 | 103 | 96 | 116 | 108 | 84 | 117 | 84 | 78 |
| July 2019 – June 2020 | 126 | 111 | 111 | 117 | 94 | 102 | 126 | 119 | 111 | 104 | 90 | 60 |
| year-on-year (%) | 101.61 | 105.71 | 102.78 | 100.00 | 91.26 | 106.25 | 108.62 | 110.19 | 132.14 | 88.89 | 107.14 | 76.92 |
| **Chemotherapy** |  |  |  |  |  |  |  |  |  |  |  |  |
| July 2018 – June 2019 | 367 | 303 | 281 | 320 | 297 | 232 | 351 | 336 | 299 | 307 | 302 | 211 |
| July 2019 – June 2020 | 346 | 304 | 301 | 321 | 291 | 273 | 343 | 284 | 277 | 265 | 274 | 185 |
| year-on-year (%) | 94.28 | 100.33 | 107.12 | 100.31 | 97.98 | 117.67 | 97.72 | 84.52 | 92.64 | 86.32 | 90.73 | 87.68 |
| **Uterine Cancer** |  |  |  |  |  |  |  |  |  |  |  |  |
| **All** |  |  |  |  |  |  |  |  |  |  |  |  |
| July 2018 – June 2019 | 668 | 653 | 595 | 655 | 614 | 516 | 673 | 581 | 590 | 583 | 616 | 493 |
| July 2019 – June 2020 | 757 | 673 | 610 | 691 | 603 | 547 | 647 | 572 | 654 | 631 | 535 | 461 |
| year-on-year (%) | 113.32 | 103.06 | 102.52 | 105.50 | 98.21 | 106.01 | 96.14 | 98.45 | 110.85 | 108.23 | 86.85 | 93.51 |
| **Surgery** |  |  |  |  |  |  |  |  |  |  |  |  |
| July 2018 – June 2019 | 182 | 163 | 140 | 186 | 162 | 129 | 200 | 187 | 166 | 169 | 176 | 145 |
| July 2019 – June 2020 | 229 | 185 | 169 | 183 | 179 | 155 | 166 | 150 | 208 | 201 | 166 | 105 |
| year-on-year (%) | 125.82 | 113.50 | 120.71 | 98.39 | 110.49 | 120.16 | 83.00 | 80.21 | 125.30 | 118.93 | 94.32 | 72.41 |
| **Chemotherapy** |  |  |  |  |  |  |  |  |  |  |  |  |
| July 2018 – June 2019 | 360 | 366 | 331 | 360 | 333 | 286 | 367 | 311 | 309 | 314 | 320 | 285 |
| July 2019 – June 2020 | 392 | 349 | 324 | 388 | 322 | 279 | 367 | 315 | 335 | 329 | 273 | 275 |
| year-on-year (%) | 108.89 | 95.36 | 97.89 | 107.78 | 96.70 | 97.55 | 100.00 | 101.29 | 108.41 | 104.78 | 85.31 | 96.49 |
| **Gallbladder & Biliary tract Cancer** |  |  |  |  |  |  |  |  |  |  |  |  |
| **All** |  |  |  |  |  |  |  |  |  |  |  |  |
| July 2018 – June 2019 | 584 | 623 | 524 | 573 | 561 | 559 | 602 | 458 | 597 | 493 | 510 | 312 |
| July 2019 – June 2020 | 642 | 547 | 586 | 583 | 519 | 546 | 599 | 540 | 549 | 538 | 516 | 295 |
| year-on-year (%) | 109.93 | 87.80 | 111.83 | 101.75 | 92.51 | 97.67 | 99.50 | 117.90 | 91.96 | 109.13 | 101.18 | 94.55 |
| **Surgery** |  |  |  |  |  |  |  |  |  |  |  |  |
| July 2018 – June 2019 | 36 | 34 | 27 | 28 | 36 | 31 | 50 | 32 | 39 | 31 | 28 | 16 |
| July 2019 – June 2020 | 36 | 30 | 36 | 46 | 22 | 24 | 28 | 30 | 24 | 33 | 28 | 9 |
| year-on-year (%) | 100.00 | 88.24 | 133.33 | 164.29 | 61.11 | 77.42 | 56.00 | 93.75 | 61.54 | 106.45 | 100.00 | 56.25 |
| **Chemotherapy** |  |  |  |  |  |  |  |  |  |  |  |  |
| July 2018 – June 2019 | 89 | 113 | 94 | 99 | 93 | 80 | 76 | 62 | 78 | 61 | 86 | 59 |
| July 2019 – June 2020 | 109 | 79 | 82 | 102 | 75 | 58 | 109 | 72 | 72 | 74 | 79 | 55 |
| year-on-year (%) | 122.47 | 69.91 | 87.23 | 103.03 | 80.65 | 72.50 | 143.42 | 116.13 | 92.31 | 121.31 | 91.86 | 93.22 |

Abbreviations; TUR-BT: Transurethral resection of bladder tumor, RFA: Radiofrequency ablation, TACE: Transcatheter arterial chemoembolization, TUR-P: Transurethral resection of prostate

We conducted ITS analysis for the number of cases for the total study population and each type of cancer diagnosis. The results indicated that the COVID-19 pandemic had a negative impact on the number of cases for most types of cancer (Table4). In particular, the number of cases with gastric and esophageal cancer was affected more. The result of ITS analysis for the overall study population is described in Figure1. In the ITS analysis, the number of cases per month was counted on the basis of the date of discharge. These results are shown in Supplementary Table2.

**Table4.**
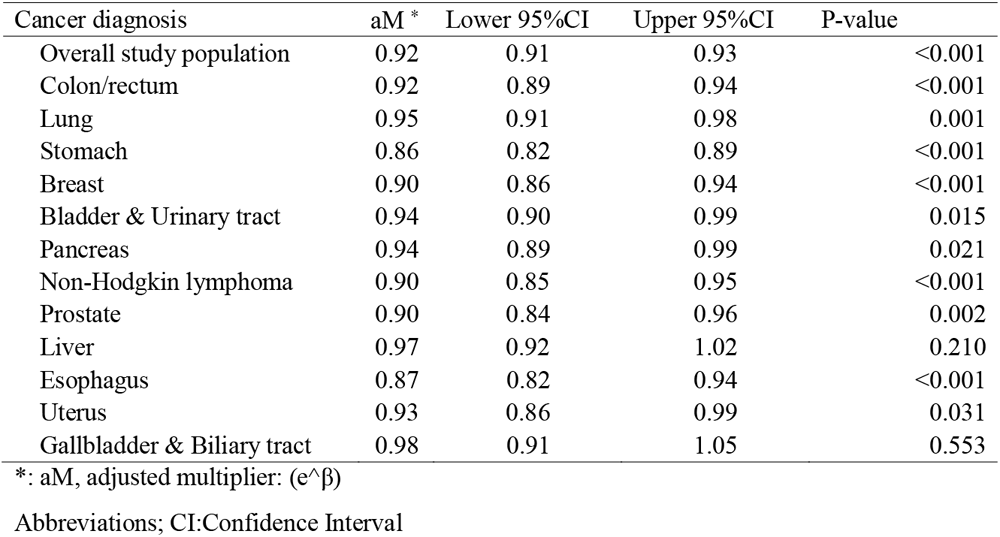
Interrupted time series analysis for total admissions cases of each cancer diagnois

| Cancer diagnosis | aM * | Lower 95%CI | Upper 95%CI | P-value |
| --- | --- | --- | --- | --- |
| Overall study population | 0.92 | 0.91 | 0.93 | <0.001 |
| Colon/rectum | 0.92 | 0.89 | 0.94 | <0.001 |
| Lung | 0.95 | 0.91 | 0.98 | 0.001 |
| Stomach | 0.86 | 0.82 | 0.89 | <0.001 |
| Breast | 0.90 | 0.86 | 0.94 | <0.001 |
| Bladder & Urinary tract | 0.94 | 0.90 | 0.99 | 0.015 |
| Pancreas | 0.94 | 0.89 | 0.99 | 0.021 |
| Non-Hodgkin lymphoma | 0.90 | 0.85 | 0.95 | <0.001 |
| Prostate | 0.90 | 0.84 | 0.96 | 0.002 |
| Liver | 0.97 | 0.92 | 1.02 | 0.210 |
| Esophagus | 0.87 | 0.82 | 0.94 | <0.001 |
| Uterus | 0.93 | 0.86 | 0.99 | 0.031 |
| Gallbladder & Biliary tract | 0.98 | 0.91 | 1.05 | 0.553 |

\*: aM, adjusted multiplier: ( $e^{\beta}$ )
Abbreviations; CI:Confidence Interval

**Figure1.**
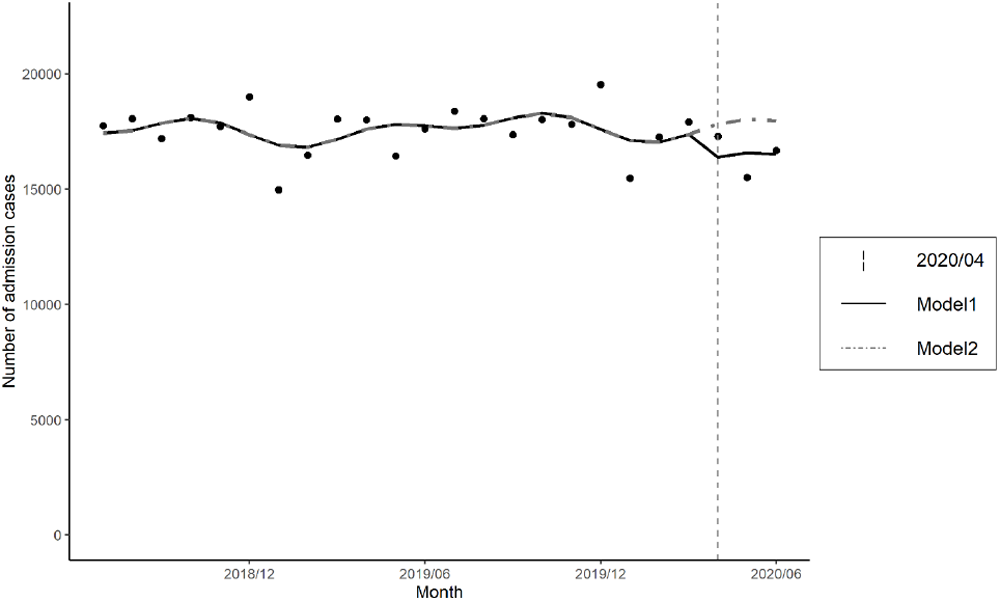
Interrupted time series with level change regression model for the overall study population. Model1: Predicted trend based on the seasonally adjusted regression model during the COVID-19outbreak. Model2: Counterfactual scenario (COVID-19 outbreak did not occur).

## DISCUSSION

Regarding cancer practice during the COVID-19 pandemic, one of the main concerns is it might delay the diagnosis and treatment of cancer. Our study indicates that cancer care needed to hospitalize was affected by the COVID-19 outbreak in many cancer types. As the number of patients infected with COVID-19 increased, the Japanese government declared a state of emergency, and as a result, the number of admission cases with most types of cancer has decreased, with the lowest number in May 2020. Our study also showed that the impact differed according to the types of cancer. For cases with non-Hodgkin lymphoma, prostate, pancreas or gallbladder & biliary tract cancer, the reduction in the number of hospitalizations was less than 10% compared with other types of cancer at May, 2020. These results might have been influenced by academic recommendations or by the biological grade of the cancer itself. Pancreatic cancer was reported with a tumor doubling time of 159 days, and a cellular doubling time of 56 hours was reported in gallbladder cancer. Both cancers were known for poor overall survival [19-21]. Although there are no recommendations for non-Hodgkin’s lymphoma from a Japanese academic society, the European Society for Medical Oncology (ESMO) defined treatment of non-Hodgkin’s lymphoma as high or medium priority [5]. The American Society of Hematology also recommended that chemotherapies such as R-CHOP (rituximab plus cyclophosphamide, doxorubicin, vincristine, and prednisone) be continued [22].

In contrast, cases of lung, colorectal, gastric, and esophageal cancer decreased substantially. These results might be related to the route of infection of COVID-19. The basic infection route of COVID-19 is considered to be droplets and contacts infection [23]. Also, there are concerns about the possibility of aerosol transmission, which may have led to a substantial reduction in the number of hospitalized patients with these types of cancer [24]. Bronchoscopy and gastrointestinal endoscopy play important roles in the diagnosis of these carcinomas. However, concerns about aerosolized and fecal transmission to health care workers caused academic societies to recommend that nonurgent testing be considered for postponement [25-28]. Therefore, unlike for other types of cancer, a decrease in the number of hospitalized patients diagnosed with lung, colorectal, gastric, or esophageal cancer might indicate a decrease in the number of examinations for diagnosis rather than a delay in treatments for diagnosed patients.

This hypothesis can be inferred from Table3, which shows a similar decrease in the number of admissions with surgery or chemotherapy despite the fact that delaying treatment such as surgery and chemotherapy is not recommended. The number of cases with surgery or chemotherapy for other types of cancer did not considerably decrease. For breast cancer cases, total admissions cases were reduced by more than 10%. The number of cases with chemotherapy was also reduced by 10%, while the number of cases with surgery was reduced by less than 10%. In this regard, it is possible that the chemotherapy regimen was changed to oral drug, or the dosing interval was extended, as described in the ESMO guidelines, which may have contributed to a reduction in the overall number of hospital admissions [5]. Uterine cancer cases showed a similar trend, with the ESMO guidelines regarding second-line chemotherapy as a low priority, which might explain the decline in the number of chemotherapy cases [5].

This study had several limitations. We evaluated the impact of COVID-19 on cancer care for inpatients, but not for outpatient. Events related to outpatient cancer treatment include postponing outpatient testing if they were suspected of having cancer or postponing chemotherapy in outpatients. Because chemotherapy is often administered as an outpatient procedure, it is unclear how outpatients with cancer were affected. It is also unclear how a decrease in the number of hospitalized patients is associated with cancer outcomes. In fact, the decline in the number of admission cases treated with surgery or chemotherapy was small except for some types of cancer patients, so it might have little impact on long-term prognosis. The long-term prognosis should be carefully monitored. In addition, due to the characteristics of DPC data, the number of patients who were admitted in June decreased significantly in both 2019 and 2020. Many hospitalized patients in June may still have been admitted; therefore, no DPC data are not generated without discharge, making it difficult to assess the impact of the COVID-19 pandemic in June 2020.

In conclusion, our study indicated the COVID-19 outbreak might cause a decrease in the number of admission cases with cancer, and cases that received treatments such as surgery or chemotherapy were not largely affected except for some types of cancer.

## Data Availability

The datasets generated during and/or analyzed during the present study are available from the corresponding author on reasonable request.

## Author contributions

Conceptualization: All authors

Methodology: HI, YA, DT and TM

Software: HI, JS, DT, and TM

Validation: All authors

Formal analysis: HI, YA, and JS

Investigation: All authors

Resources: SK and YI

Data Curation: JS, and SK

Writing – original draft preparation: HI

Writing – review and editing: All authors

Visualization: HI

Supervision: YI

Project administration: YI

Funding acquisition: YI

## Acknowledgments

The present study was supported by JSPS KAKENHI Grant numbers JP19H01075 from the Japan Society for the Promotion of Science, Health Labour Sciences Research Grants from the Ministry of Health, Labour and Welfare, Japan, Grant Numbers 20HA2003 and by GAP Fund Program of Kyoto University, Type B (2020) to Yuichi Imanaka.

