## Supplementary_Materials for "The impact of the coronavirus disease 2019 (COVID-19) outbreak on cancer practice in Japan: using an administrative database"

**Contents**

Supplementary Figure1. Line graph of admission cases for each cancer diagnosis

Supplementary Table1. Baseline characteristics of study population for each cancer diagnosis

Supplementary Table2. Number of monthly admission cases for each treatment for each diagnosis of cancer based on the date of discharge

**Supplementary Figure1. Line graph of admission cases for each cancer diagnosis**

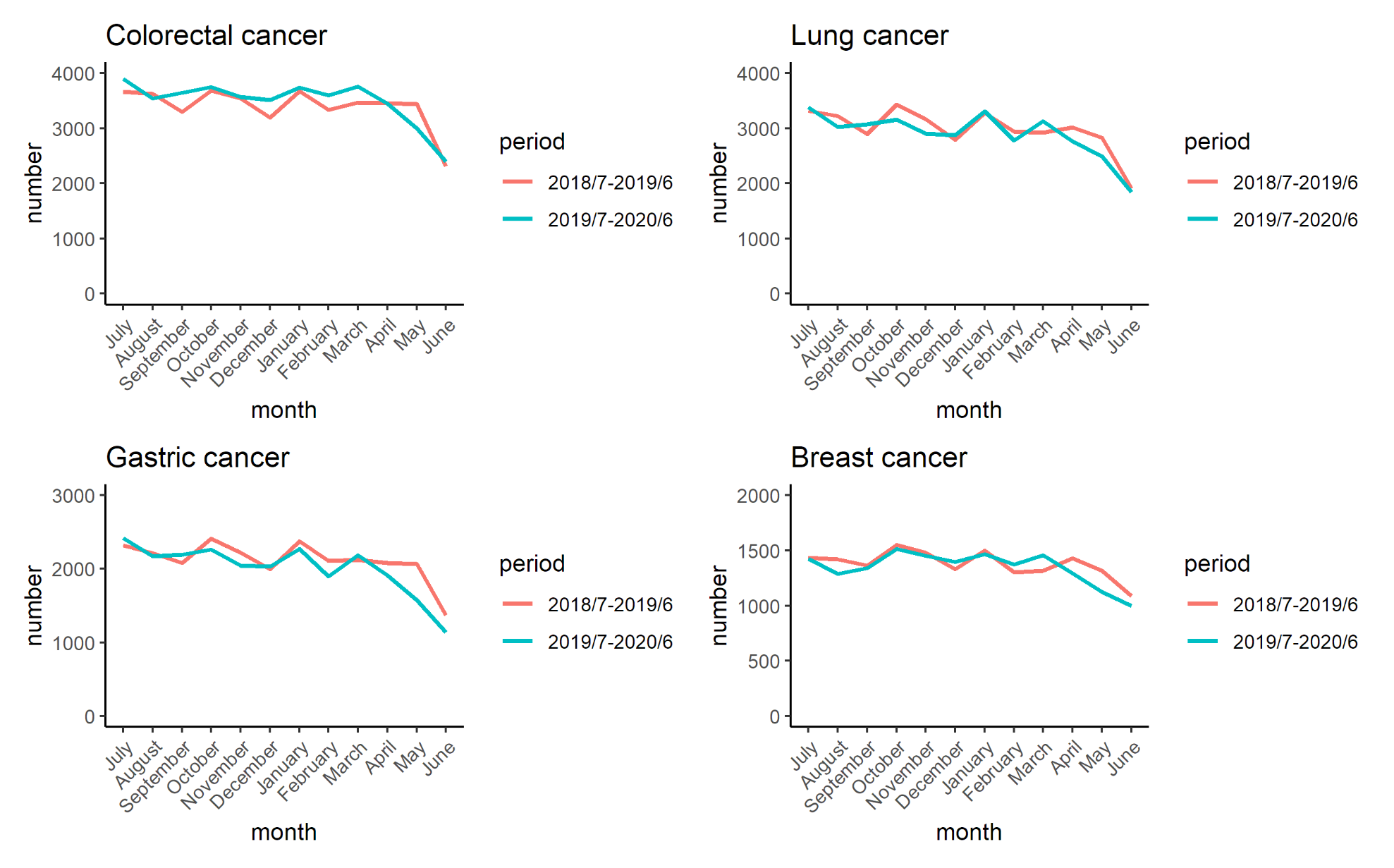

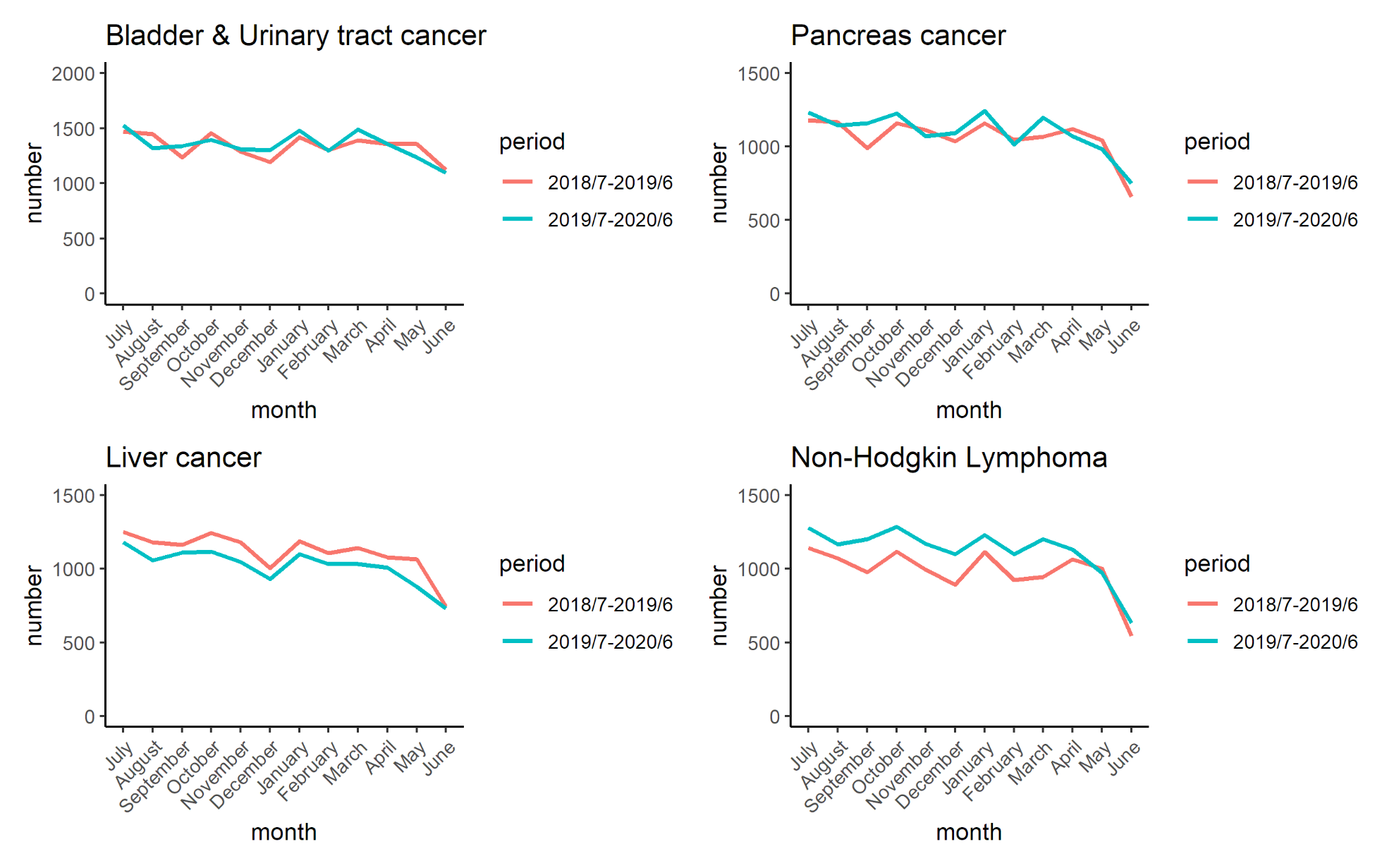

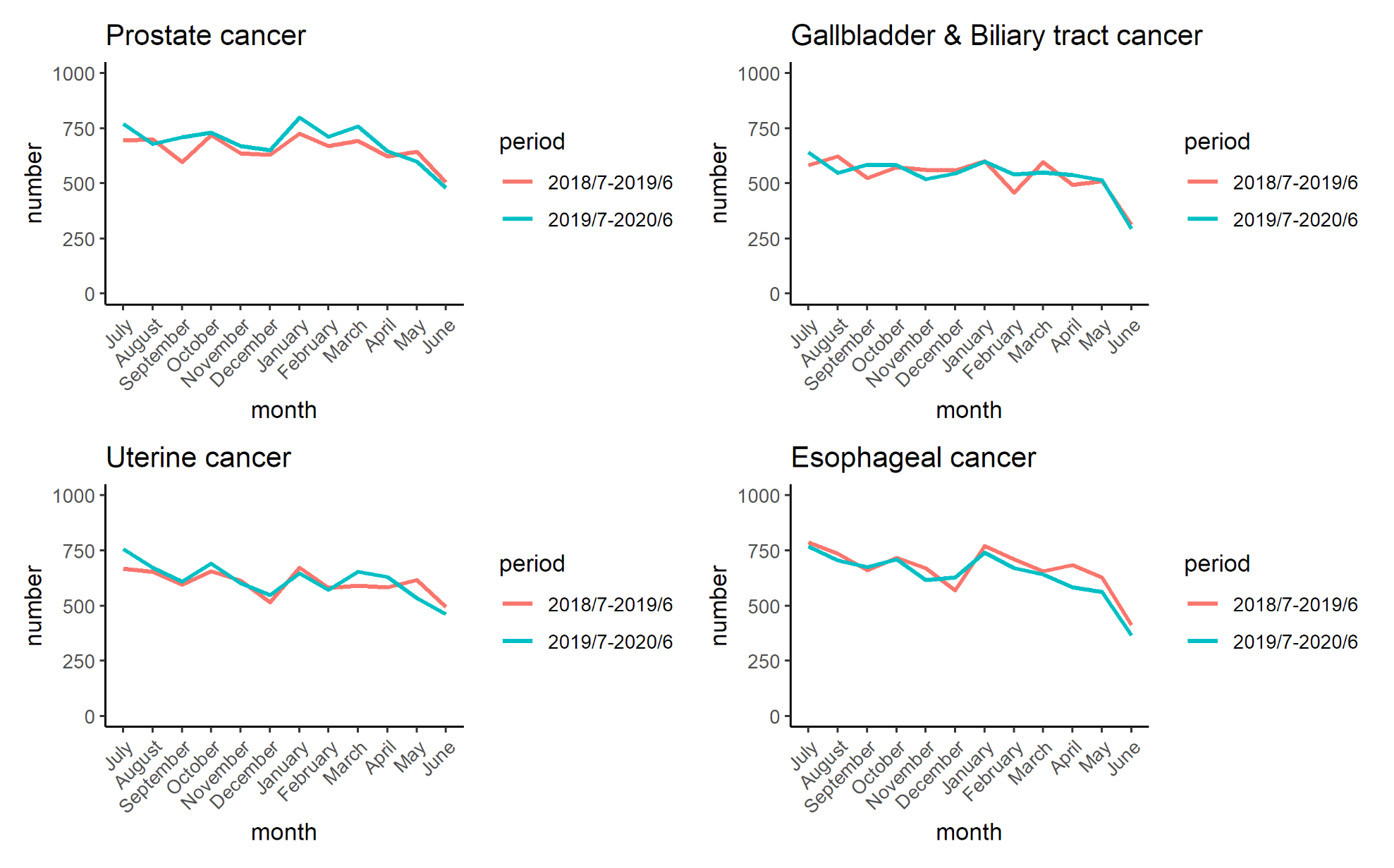

**Supplementary Table1. Baseline characteristics of study population for each cancer diagnosis**

| Characteristic | Colorectal  (n = 82,541) | | Lung  (n = 70,455) | | Stomach  (n = 49,504) | | Breast  (n = 32,662) | | Bladder & Urinary tract  (n = 32,186) | | Pancreas  (n = 25,948) | |
| --- | --- | --- | --- | --- | --- | --- | --- | --- | --- | --- | --- | --- |
| **Age** |  |  |  |  |  |  |  |  |  |  |  |  |
| Median (IQR) | 71 | (65, 78) | 72 | (67, 78) | 73 | (67, 80) | 63 | (51, 72) | 75 | (68, 81) | 72 | (65, 78) |
| **Age category** |  |  |  |  |  |  |  |  |  |  |  |  |
| 75 years ≥ | 31,120 | 37.7% | 27,485 | 39.0% | 22,607 | 45.7% | 6,249 | 19.1% | 16,553 | 51.4% | 10,381 | 40.0% |
| 65 – 74 years | 30,857 | 37.4% | 29,400 | 41.7% | 18,205 | 36.8% | 8,775 | 26.9% | 10,880 | 33.8% | 9,490 | 36.6% |
| 64 years ≤ | 20,564 | 24.9% | 13,570 | 19.3% | 8,692 | 17.6% | 17,638 | 54.0% | 4,773 | 14.8% | 6,077 | 23.4% |
| **Sex** |  |  |  |  |  |  |  |  |  |  |  |  |
| Males | 47,222 | 57.2% | 49,007 | 69.6% | 34,887 | 70.5% | 156 | 0.5% | 25,156 | 78.2% | 14,019 | 54.0% |
| Females | 35,319 | 42.8% | 21,448 | 30.4% | 14,617 | 29.5% | 32,506 | 99.5% | 7,030 | 21.8% | 11,929 | 46.0% |
| **Primary/Recurrent** |  |  |  |  |  |  |  |  |  |  |  |  |
| Primary | 62,242 | 75.4% | 49,072 | 69.7% | 39,994 | 80.8% | 27,124 | 83.0% | 19,969 | 62.0% | 19,128 | 73.7% |
| Recurrent | 18,307 | 22.2% | 19,056 | 27.0% | 8,018 | 16.2% | 4,959 | 15.2% | 11,825 | 36.7% | 5,544 | 21.4% |
| Unknown | 1,992 | 2.4% | 2,327 | 3.3% | 1,492 | 3.0% | 579 | 1.8% | 392 | 1.2% | 1,276 | 4.9% |
| **Purpose of admission** |  |  |  |  |  |  |  |  |  |  |  |  |
| Special purpose | 80,961 | 98.1% | 65,447 | 92.9% | 49,012 | 99.0% | 32,616 | 99.9% | 32,065 | 99.6% | 24,971 | 96.2% |
| Examination | 1,580 | 1.9% | 5,008 | 7.1% | 492 | 1.0% | 46 | 0.1% | 121 | 0.4% | 977 | 3.8% |
| **Anti-cancer treatment** |  |  |  |  |  |  |  |  |  |  |  |  |
| Any surgery | 30,762 | 37.4% | 10,442 | 14.8% | 23,672 | 47.8% | 19,118 | 58.5% | 21,520 | 66.9% | 2,320 | 8.9% |
| Any chemotherapy | 26,958 | 32.7% | 36,604 | 52.0% | 11,038 | 22.4% | 8,593 | 26.0% | 10,389 | 33.1% | 8,939 | 34.4% |
| Length of hospital stay, median (IQR) | 9 | (4, 17) | 10 | (5, 18) | 10 | (6, 17) | 7 | (5, 18) | 7 | (5, 10) | 11 | (5, 22) |
| In-hospital mortality | 4,961 | 8.7% | 8,445 | 12.0% | 4,564 | 9.2% | 1,352 | 4.1% | 1,047 | 3.3% | 4,765 | 18.4% |

| Characteristic | Non-Hodgkin lymphoma  (n = 25,273) | | Liver  (n = 25,588) | | Prostate  (n = 16,045) | | Esophagus  (n = 15,668) | | Uterus  (n = 14,618) | | Gallbladder & Biliary tract  (n = 12,856) | |
| --- | --- | --- | --- | --- | --- | --- | --- | --- | --- | --- | --- | --- |
| **Age** |  |  |  |  |  |  |  |  |  |  |  |  |
| Median (IQR) | 73 | (66, 80) | 75 | (68, 81) | 73 | (68, 78) | 71 | (65, 77) | 60 | (50, 70) | 77 | (70, 83) |
| **Age category** |  |  |  |  |  |  |  |  |  |  |  |  |
| 75 years ≥ | 11,778 | 40.0% | 13,154 | 40.0% | 6,721 | 41.9% | 5,574 | 35..6% | 2,129 | 14.6% | 7,768 | 60.4% |
| 65 – 74 years | 8,063 | 36.6% | 8,646 | 36.6% | 7,286 | 45.4% | 6,579 | 42.0% | 3,734 | 25.5% | 3,716 | 28.9% |
| 64 years ≤ | 5,432 | 23.4% | 3,788 | 23.4% | 2,038 | 12.7% | 3,515 | 22.4% | 8,755 | 59.9% | 1,372 | 10.7% |
| **Sex** |  |  |  |  |  |  |  |  |  |  |  |  |
| Males | 13,625 | 54.0% | 18,685 | 54.0% | 16,045 | 100.0% | 12,889 | 82.3% | 0 | 0.0% | 7,654 | 59.5% |
| Females | 11,648 | 46.0% | 6,903 | 46.0% | 0 | 0.0% | 2,779 | 17.7% | 14,618 | 100.0% | 5,202 | 40.5% |
| **Primary/Recurrent** |  |  |  |  |  |  |  |  |  |  |  |  |
| Primary | 17,294 | 68.4% | 11,512 | 45.0% | 12,475 | 77.8% | 11,822 | 75.5% | 10,701 | 73.2% | 9,635 | 74.9% |
| Recurrent | 7,657 | 30.3% | 13,365 | 52.2% | 3,179 | 19.8% | 3,336 | 21.2% | 3,543 | 24.2% | 2,712 | 21.1% |
| Unknown | 322 | 1.3% | 711 | 2.8% | 391 | 2.4% | 510 | 3.3% | 374 | 2.6% | 509 | 4.0% |
| **Purpose of admission** |  |  |  |  |  |  |  |  |  |  |  |  |
| Special purpose | 24,965 | 98.8% | 25,074 | 98.0% | 14,145 | 88.2% | 15,569 | 99.4% | 14,586 | 99.8% | 12,694 | 98.7% |
| Examination | 308 | 1.2% | 514 | 2.0% | 1,900 | 11.8% | 99 | 0.6% | 32 | 0.2% | 162 | 1.3% |
| **Anti-cancer treatment** |  |  |  |  |  |  |  |  |  |  |  |  |
| Any surgery | 0 | 0.0% | 13,814 | 54.0% | 5,800 | 36.1% | 3,532 | 22.5% | 4,101 | 28.1% | 734 | 5.7% |
| Any chemotherapy | 20,094 | 79.5% | 3,555 | 13.9% | 3,946 | 24.6% | 7,070 | 45.1% | 7,890 | 54.0% | 1,956 | 15.2% |
| Length of hospital stay, median (IQR) | 16 | (9, 23) | 9 | (6, 15) | 10 | (4, 14) | 9 | (7, 19) | 6 | (3, 12) | 6 | (3, 12) |
| In-hospital mortality | 1,708 | 6.8% | 3,019 | 11.8% | 971 | 6.1% | 1,428 | 9.1% | 769 | 5.3% | 1,986 | 15.4% |

Abbreviations; IQR: Interquartile range

**Supplementary Table2. Number of monthly admission cases for each treatment for each diagnosis of cancer based on the date of discharge**

|  | | Jul | | Aug | | Sep | | Oct | | Nov | | Dec | | Jan | | Feb | | Mar | | Apr | | May | | Jun |
| --- | --- | --- | --- | --- | --- | --- | --- | --- | --- | --- | --- | --- | --- | --- | --- | --- | --- | --- | --- | --- | --- | --- | --- | --- |
| **All Cancers** |  | |  | |  | |  | |  | |  | |  | |  | |  | |  | |  | |  | |
| July 2018 – June 2019 | | 21,741 | | 22,274 | | 21,120 | | 22,224 | | 21,710 | | 23,212 | | 18,255 | | 20,120 | | 21,995 | | 21,970 | | 20,164 | | 21,568 |
| July 2019 – June 2020 | | 22,603 | | 22,130 | | 21,246 | | 22,085 | | 21,728 | | 23,946 | | 18,968 | | 21,114 | | 21,834 | | 21,184 | | 19,122 | | 20,478 |
| year-on-year (%) | | 103.96 | | 99.35 | | 100.60 | | 99.37 | | 100.08 | | 103.16 | | 103.91 | | 104.94 | | 99.27 | | 96.42 | | 94.83 | | 94.95 |
| **Overall study population** | |  | |  | |  | |  | |  | |  | |  | |  | |  | |  | |  | |  |
| July 2018 – June 2019 | | 17,745 | | 18,052 | | 17,202 | | 18,114 | | 17,716 | | 18,998 | | 14,968 | | 16,470 | | 18,047 | | 18,016 | | 16,430 | | 17,624 |
| July 2019 – June 2020 | | 18,390 | | 18,051 | | 17,363 | | 18,023 | | 17,813 | | 19,530 | | 15,477 | | 17,261 | | 17,909 | | 17,288 | | 15,503 | | 16,681 |
| year-on-year (%) | | 103.63 | | 99.99 | | 100.94 | | 99.50 | | 100.55 | | 102.80 | | 103.40 | | 104.80 | | 99.24 | | 95.96 | | 94.36 | | 94.65 |
| **Colorectal Cancer** | |  | |  | |  | |  | |  | |  | |  | |  | |  | |  | |  | |  |
| **All** | |  | |  | |  | |  | |  | |  | |  | |  | |  | |  | |  | |  |
| July 2018 – June 2019 | | 3,490 | | 3,653 | | 3,435 | | 3,610 | | 3,514 | | 3,787 | | 3,132 | | 3,322 | | 3,662 | | 3,615 | | 3,406 | | 3,628 |
| July 2019 – June 2020 | | 3,711 | | 3,764 | | 3,542 | | 3,669 | | 3,701 | | 3,993 | | 3,222 | | 3,646 | | 3,806 | | 3,633 | | 3,232 | | 3,499 |
| year-on-year (%) | | 106.33 | | 103.04 | | 103.11 | | 101.63 | | 105.32 | | 105.44 | | 102.87 | | 109.75 | | 103.93 | | 100.50 | | 94.89 | | 96.44 |
| **Proportion of All**  **Colorectal Cancer to**  **All Cancers** | |  | |  | |  | |  | |  | |  | |  | |  | |  | |  | |  | |  |
| July 2018 – June 2019 | | 0.16 | | 0.16 | | 0.16 | | 0.16 | | 0.16 | | 0.16 | | 0.17 | | 0.17 | | 0.17 | | 0.16 | | 0.17 | | 0.17 |
| July 2019 – June 2020 | | 0.16 | | 0.17 | | 0.17 | | 0.17 | | 0.17 | | 0.17 | | 0.17 | | 0.17 | | 0.17 | | 0.17 | | 0.17 | | 0.17 |
| Ratio of proportion* | | 1.02 | | 1.04 | | 1.03 | | 1.02 | | 1.05 | | 1.02 | | 0.99 | | 1.05 | | 1.05 | | 1.04 | | 1.00 | | 1.02 |
| **Surgery** | |  | |  | |  | |  | |  | |  | |  | |  | |  | |  | |  | |  |
| July 2018 – June 2019 | | 1,092 | | 1,186 | | 1,058 | | 1,133 | | 1,215 | | 1,246 | | 953 | | 1,090 | | 1,152 | | 1,181 | | 1,063 | | 1,116 |
| July 2019 – June 2020 | | 1,183 | | 1,181 | | 1,145 | | 1,194 | | 1,180 | | 1,295 | | 951 | | 1,144 | | 1,276 | | 1,142 | | 1,058 | | 1,019 |
| year-on-year (%) | | 108.33 | | 99.58 | | 108.22 | | 105.38 | | 97.12 | | 103.93 | | 99.79 | | 104.95 | | 110.76 | | 96.70 | | 99.53 | | 91.31 |
| **Endoscopic resection** | |  | |  | |  | |  | |  | |  | |  | |  | |  | |  | |  | |  |
| July 2018 – June 2019 | | 188 | | 180 | | 169 | | 193 | | 181 | | 220 | | 182 | | 212 | | 207 | | 212 | | 159 | | 229 |
| July 2019 – June 2020 | | 220 | | 217 | | 228 | | 238 | | 230 | | 267 | | 180 | | 250 | | 263 | | 269 | | 184 | | 233 |
| year-on-year (%) | | 117.02 | | 120.56 | | 134.91 | | 123.32 | | 127.07 | | 121.36 | | 98.90 | | 117.92 | | 127.05 | | 126.89 | | 115.72 | | 101.75 |
| **Chemotherapy** | |  | |  | |  | |  | |  | |  | |  | |  | |  | |  | |  | |  |
| July 2018 – June 2019 | | 1,141 | | 1,182 | | 1,141 | | 1,106 | | 1,076 | | 1,125 | | 1,053 | | 1,051 | | 1,119 | | 1,124 | | 1,115 | | 1,175 |
| July 2019 – June 2020 | | 1,160 | | 1,222 | | 1,054 | | 1,143 | | 1,199 | | 1,193 | | 1,139 | | 1,170 | | 1,143 | | 1,252 | | 1,138 | | 1,125 |
| year-on-year (%) | | 101.67 | | 103.38 | | 92.38 | | 103.35 | | 111.43 | | 106.04 | | 108.17 | | 111.32 | | 102.14 | | 111.39 | | 102.06 | | 95.74 |
| **Lung Cancer** | |  | |  | |  | |  | |  | |  | |  | |  | |  | |  | |  | |  |
| **All** | |  | |  | |  | |  | |  | |  | |  | |  | |  | |  | |  | |  |
| July 2018 – June 2019 | | 3,133 | | 3,180 | | 3,047 | | 3,245 | | 3,289 | | 3,369 | | 2,698 | | 2,910 | | 3,171 | | 3,159 | | 2,821 | | 3,007 |
| July 2019 – June 2020 | | 3,169 | | 3,159 | | 2,987 | | 3,113 | | 3,078 | | 3,388 | | 2,703 | | 2,911 | | 3,026 | | 2,926 | | 2,722 | | 2,855 |
| year-on-year (%) | | 101.15 | | 99.34 | | 98.03 | | 95.93 | | 93.58 | | 100.56 | | 100.19 | | 100.03 | | 95.43 | | 92.62 | | 96.49 | | 94.95 |
| **Proportion of All Lung**  **Cancer to All Cancers** | |  | |  | |  | |  | |  | |  | |  | |  | |  | |  | |  | |  |
| July 2018 – June 2019 | | 0.14 | | 0.14 | | 0.14 | | 0.15 | | 0.15 | | 0.15 | | 0.15 | | 0.14 | | 0.14 | | 0.14 | | 0.14 | | 0.14 |
| July 2019 – June 2020 | | 0.14 | | 0.14 | | 0.14 | | 0.14 | | 0.14 | | 0.14 | | 0.14 | | 0.14 | | 0.14 | | 0.14 | | 0.14 | | 0.14 |
| Rate of proportion | | 0.97 | | 1.00 | | 0.97 | | 0.97 | | 0.94 | | 0.97 | | 0.96 | | 0.95 | | 0.96 | | 0.96 | | 1.02 | | 1.00 |
| **Surgery** | |  | |  | |  | |  | |  | |  | |  | |  | |  | |  | |  | |  |
| July 2018 – June 2019 | | 430 | | 446 | | 443 | | 462 | | 495 | | 504 | | 391 | | 475 | | 457 | | 484 | | 364 | | 481 |
| July 2019 – June 2020 | | 469 | | 435 | | 434 | | 495 | | 437 | | 559 | | 385 | | 462 | | 478 | | 412 | | 364 | | 386 |
| year-on-year (%) | | 109.07 | | 97.53 | | 97.97 | | 107.14 | | 88.28 | | 110.91 | | 98.47 | | 97.26 | | 104.60 | | 85.12 | | 100.00 | | 80.25 |
| **Chemotherapy** | |  | |  | |  | |  | |  | |  | |  | |  | |  | |  | |  | |  |
| July 2018 – June 2019 | | 1,600 | | 1,623 | | 1,554 | | 1,676 | | 1,755 | | 1,817 | | 1,367 | | 1,510 | | 1,700 | | 1,647 | | 1,524 | | 1,541 |
| July 2019 – June 2020 | | 1,617 | | 1,676 | | 1,511 | | 1,556 | | 1,576 | | 1,708 | | 1,398 | | 1,523 | | 1,576 | | 1,539 | | 1,450 | | 1,501 |
| year-on-year (%) | | 101.06 | | 103.27 | | 97.23 | | 92.84 | | 89.80 | | 94.00 | | 102.27 | | 100.86 | | 92.71 | | 93.44 | | 95.14 | | 97.40 |
| **Gastric Cancer** | |  | |  | |  | |  | |  | |  | |  | |  | |  | |  | |  | |  |
| **All** | |  | |  | |  | |  | |  | |  | |  | |  | |  | |  | |  | |  |
| July 2018 – June 2019 | | 2,251 | | 2,225 | | 2,145 | | 2,353 | | 2,287 | | 2,412 | | 1,850 | | 2,102 | | 2,370 | | 2,257 | | 1,986 | | 2,174 |
| July 2019 – June 2020 | | 2,326 | | 2,188 | | 2,222 | | 2,215 | | 2,105 | | 2,454 | | 1,919 | | 1,980 | | 2,190 | | 2,065 | | 1,663 | | 1,785 |
| year-on-year (%) | | 103.33 | | 98.34 | | 103.59 | | 94.14 | | 92.04 | | 101.74 | | 103.73 | | 94.20 | | 92.41 | | 91.49 | | 83.74 | | 82.11 |
| **Proportion of All Gastric**  **Cancer to All Cancers** | |  | |  | |  | |  | |  | |  | |  | |  | |  | |  | |  | |  |
| July 2018 – June 2019 | | 0.10 | | 0.10 | | 0.10 | | 0.11 | | 0.11 | | 0.10 | | 0.10 | | 0.10 | | 0.11 | | 0.10 | | 0.10 | | 0.10 |
| July 2019 – June 2020 | | 0.10 | | 0.10 | | 0.10 | | 0.10 | | 0.10 | | 0.10 | | 0.10 | | 0.09 | | 0.10 | | 0.10 | | 0.09 | | 0.09 |
| Ratio of proportion | | 0.99 | | 0.99 | | 1.03 | | 0.95 | | 0.92 | | 0.99 | | 1.00 | | 0.90 | | 0.93 | | 0.95 | | 0.88 | | 0.86 |
| **Surgery** | |  | |  | |  | |  | |  | |  | |  | |  | |  | |  | |  | |  |
| July 2018 – June 2019 | | 502 | | 505 | | 452 | | 518 | | 515 | | 518 | | 384 | | 462 | | 538 | | 448 | | 381 | | 435 |
| July 2019 – June 2020 | | 483 | | 474 | | 480 | | 504 | | 442 | | 574 | | 366 | | 430 | | 439 | | 462 | | 381 | | 354 |
| year-on-year (%) | | 96.22 | | 93.86 | | 106.19 | | 97.30 | | 85.83 | | 110.81 | | 95.31 | | 93.07 | | 81.60 | | 103.13 | | 100.00 | | 81.38 |
| **Endoscopic resection** | |  | |  | |  | |  | |  | |  | |  | |  | |  | |  | |  | |  |
| July 2018 – June 2019 | | 590 | | 549 | | 553 | | 592 | | 594 | | 631 | | 451 | | 531 | | 577 | | 602 | | 444 | | 567 |
| July 2019 – June 2020 | | 673 | | 600 | | 606 | | 590 | | 551 | | 721 | | 509 | | 599 | | 673 | | 567 | | 376 | | 426 |
| year-on-year (%) | | 114.07 | | 109.29 | | 109.58 | | 99.66 | | 92.76 | | 114.26 | | 112.86 | | 112.81 | | 116.64 | | 94.19 | | 84.68 | | 75.13 |
| **Chemotherapy** | |  | |  | |  | |  | |  | |  | |  | |  | |  | |  | |  | |  |
| July 2018 – June 2019 | | 506 | | 518 | | 495 | | 550 | | 519 | | 522 | | 486 | | 493 | | 570 | | 557 | | 526 | | 507 |
| July 2019 – June 2020 | | 488 | | 475 | | 461 | | 430 | | 438 | | 446 | | 441 | | 401 | | 437 | | 415 | | 351 | | 385 |
| year-on-year (%) | | 96.44 | | 91.70 | | 93.13 | | 78.18 | | 84.39 | | 85.44 | | 90.74 | | 81.34 | | 76.67 | | 74.51 | | 66.73 | | 75.94 |
| **Breast Cancer** | |  | |  | |  | |  | |  | |  | |  | |  | |  | |  | |  | |  |
| **All** | |  | |  | |  | |  | |  | |  | |  | |  | |  | |  | |  | |  |
| July 2018 – June 2019 | | 1,413 | | 1,394 | | 1,418 | | 1,461 | | 1,512 | | 1,584 | | 1,241 | | 1,311 | | 1,412 | | 1,461 | | 1,324 | | 1,367 |
| July 2019 – June 2020 | | 1,371 | | 1,371 | | 1,298 | | 1,461 | | 1,476 | | 1,637 | | 1,226 | | 1,441 | | 1,443 | | 1,363 | | 1,142 | | 1,273 |
| year-on-year (%) | | 97.03 | | 98.35 | | 91.54 | | 100.00 | | 97.62 | | 103.35 | | 98.79 | | 109.92 | | 102.20 | | 93.29 | | 86.25 | | 93.12 |
| **Proportion of All Breast   Cancer to All Cancers** | |  | |  | |  | |  | |  | |  | |  | |  | |  | |  | |  | |  |
| July 2018 – June 2019 | | 0.06 | | 0.06 | | 0.07 | | 0.07 | | 0.07 | | 0.07 | | 0.07 | | 0.07 | | 0.06 | | 0.07 | | 0.07 | | 0.06 |
| July 2019 – June 2020 | | 0.06 | | 0.06 | | 0.06 | | 0.07 | | 0.07 | | 0.07 | | 0.06 | | 0.07 | | 0.07 | | 0.06 | | 0.06 | | 0.06 |
| Ratio of proportion | | 0.93 | | 0.99 | | 0.91 | | 1.01 | | 0.98 | | 1.00 | | 0.95 | | 1.05 | | 1.03 | | 0.97 | | 0.91 | | 0.98 |
| **Surgery** | |  | |  | |  | |  | |  | |  | |  | |  | |  | |  | |  | |  |
| July 2018 – June 2019 | | 826 | | 770 | | 836 | | 889 | | 845 | | 962 | | 653 | | 745 | | 811 | | 845 | | 725 | | 790 |
| July 2019 – June 2020 | | 765 | | 815 | | 771 | | 876 | | 905 | | 1027 | | 658 | | 852 | | 883 | | 824 | | 681 | | 754 |
| year-on-year (%) | | 92.62 | | 105.84 | | 92.22 | | 98.54 | | 107.10 | | 106.76 | | 100.77 | | 114.36 | | 108.88 | | 97.51 | | 93.93 | | 95.44 |
| **Chemotherapy** | |  | |  | |  | |  | |  | |  | |  | |  | |  | |  | |  | |  |
| July 2018 – June 2019 | | 370 | | 392 | | 380 | | 359 | | 406 | | 373 | | 380 | | 363 | | 390 | | 366 | | 380 | | 367 |
| July 2019 – June 2020 | | 371 | | 335 | | 316 | | 358 | | 373 | | 379 | | 366 | | 337 | | 345 | | 332 | | 295 | | 333 |
| year-on-year (%) | | 100.27 | | 85.46 | | 83.16 | | 99.72 | | 91.87 | | 101.61 | | 96.32 | | 92.84 | | 88.46 | | 90.71 | | 77.63 | | 90.74 |
| **Bladder & Urinary tract Cancer** | |  | |  | |  | |  | |  | |  | |  | |  | |  | |  | |  | |  |
| **All** | |  | |  | |  | |  | |  | |  | |  | |  | |  | |  | |  | |  |
| July 2018 – June 2019 | | 1,381 | | 1,453 | | 1,311 | | 1,424 | | 1,336 | | 1,414 | | 1,151 | | 1,302 | | 1,452 | | 1,475 | | 1,263 | | 1,478 |
| July 2019 – June 2020 | | 1,439 | | 1,399 | | 1,335 | | 1,366 | | 1,352 | | 1,538 | | 1,218 | | 1,360 | | 1,438 | | 1,404 | | 1,281 | | 1,417 |
| year-on-year (%) | | 104.20 | | 96.28 | | 101.83 | | 95.93 | | 101.20 | | 108.77 | | 105.82 | | 104.45 | | 99.04 | | 95.19 | | 101.43 | | 95.87 |
| **Proportion of All**  **Bladder & Urinary tract**  **Cancer to All Cancers** | |  | |  | |  | |  | |  | |  | |  | |  | |  | |  | |  | |  |
| July 2018 – June 2019 | | 0.06 | | 0.07 | | 0.06 | | 0.06 | | 0.06 | | 0.06 | | 0.06 | | 0.06 | | 0.07 | | 0.07 | | 0.06 | | 0.07 |
| July 2019 – June 2020 | | 0.06 | | 0.06 | | 0.06 | | 0.06 | | 0.06 | | 0.06 | | 0.06 | | 0.06 | | 0.07 | | 0.07 | | 0.07 | | 0.07 |
| Ratio of proportion | | 1.00 | | 0.97 | | 1.01 | | 0.97 | | 1.01 | | 1.05 | | 1.02 | | 1.00 | | 1.00 | | 0.99 | | 1.07 | | 1.01 |
| **Surgery** | |  | |  | |  | |  | |  | |  | |  | |  | |  | |  | |  | |  |
| July 2018 – June 2019 | | 42 | | 39 | | 46 | | 47 | | 48 | | 58 | | 35 | | 42 | | 50 | | 59 | | 35 | | 43 |
| July 2019 – June 2020 | | 47 | | 51 | | 37 | | 57 | | 36 | | 55 | | 24 | | 44 | | 43 | | 53 | | 46 | | 48 |
| year-on-year (%) | | 111.90 | | 130.77 | | 80.43 | | 121.28 | | 75.00 | | 94.83 | | 68.57 | | 104.76 | | 86.00 | | 89.83 | | 131.43 | | 111.63 |
| **TUR-BT** | |  | |  | |  | |  | |  | |  | |  | |  | |  | |  | |  | |  |
| July 2018 – June 2019 | | 872 | | 925 | | 840 | | 893 | | 840 | | 899 | | 708 | | 843 | | 922 | | 923 | | 763 | | 961 |
| July 2019 – June 2020 | | 904 | | 873 | | 818 | | 858 | | 851 | | 1,008 | | 760 | | 871 | | 951 | | 904 | | 771 | | 885 |
| year-on-year (%) | | 103.67 | | 94.38 | | 97.38 | | 96.08 | | 101.31 | | 112.12 | | 107.34 | | 103.32 | | 103.15 | | 97.94 | | 101.05 | | 92.09 |
| **Chemotherapy** | |  | |  | |  | |  | |  | |  | |  | |  | |  | |  | |  | |  |
| July 2018 – June 2019 | | 470 | | 411 | | 428 | | 420 | | 453 | | 383 | | 438 | | 471 | | 491 | | 425 | | 451 | | 354 |
| July 2019 – June 2020 | | 482 | | 451 | | 439 | | 472 | | 442 | | 459 | | 406 | | 461 | | 441 | | 431 | | 439 | | 455 |
| year-on-year (%) | | 102.55 | | 109.73 | | 102.57 | | 112.38 | | 97.57 | | 119.84 | | 92.69 | | 97.88 | | 89.82 | | 101.41 | | 97.34 | | 128.53 |
| **Pancreas Cancer** | |  | |  | |  | |  | |  | |  | |  | |  | |  | |  | |  | |  |
| **All** | |  | |  | |  | |  | |  | |  | |  | |  | |  | |  | |  | |  |
| July 2018 – June 2019 | | 1,129 | | 1,151 | | 1,070 | | 1,137 | | 1,116 | | 1,238 | | 922 | | 1,079 | | 1,120 | | 1,172 | | 1,058 | | 1,155 |
| July 2019 – June 2020 | | 1,172 | | 1,155 | | 1,174 | | 1,209 | | 1,176 | | 1,239 | | 1,036 | | 1,125 | | 1,151 | | 1,134 | | 1,070 | | 1,164 |
| year-on-year (%) | | 103.81 | | 100.35 | | 109.72 | | 106.33 | | 105.38 | | 100.08 | | 112.36 | | 104.26 | | 102.77 | | 96.76 | | 101.13 | | 100.78 |
| **Proportion of All**  **Pancreas Cancer to All**  **Cancers** | |  | |  | |  | |  | |  | |  | |  | |  | |  | |  | |  | |  |
| July 2018 – June 2019 | | 0.05 | | 0.05 | | 0.05 | | 0.05 | | 0.05 | | 0.05 | | 0.05 | | 0.05 | | 0.05 | | 0.05 | | 0.05 | | 0.05 |
| July 2019 – June 2020 | | 0.05 | | 0.05 | | 0.06 | | 0.05 | | 0.05 | | 0.05 | | 0.05 | | 0.05 | | 0.05 | | 0.05 | | 0.06 | | 0.06 |
| Ratio of proportion | | 1.00 | | 1.01 | | 1.09 | | 1.07 | | 1.05 | | 0.97 | | 1.08 | | 0.99 | | 1.04 | | 1.00 | | 1.07 | | 1.06 |
| **Surgery** | |  | |  | |  | |  | |  | |  | |  | |  | |  | |  | |  | |  |
| July 2018 – June 2019 | | 104 | | 111 | | 101 | | 122 | | 77 | | 130 | | 76 | | 102 | | 115 | | 107 | | 89 | | 101 |
| July 2019 – June 2020 | | 121 | | 108 | | 103 | | 109 | | 112 | | 101 | | 86 | | 120 | | 112 | | 116 | | 121 | | 108 |
| year-on-year (%) | | 116.35 | | 97.30 | | 101.98 | | 89.34 | | 145.45 | | 77.69 | | 113.16 | | 117.65 | | 97.39 | | 108.41 | | 135.96 | | 106.93 |
| **Chemotherapy** | |  | |  | |  | |  | |  | |  | |  | |  | |  | |  | |  | |  |
| July 2018 – June 2019 | | 363 | | 386 | | 351 | | 391 | | 423 | | 433 | | 306 | | 371 | | 384 | | 407 | | 329 | | 412 |
| July 2019 – June 2020 | | 400 | | 361 | | 393 | | 387 | | 421 | | 432 | | 348 | | 382 | | 403 | | 372 | | 375 | | 403 |
| year-on-year (%) | | 110.19 | | 93.52 | | 111.97 | | 98.98 | | 99.53 | | 99.77 | | 113.73 | | 102.96 | | 104.95 | | 91.40 | | 113.98 | | 97.82 |
| **Non-Hodgkin Lymphoma** | |  | |  | |  | |  | |  | |  | |  | |  | |  | |  | |  | |  |
| **All** | |  | |  | |  | |  | |  | |  | |  | |  | |  | |  | |  | |  |
| July 2018 – June 2019 | | 1,115 | | 1,050 | | 1,042 | | 1,076 | | 1,041 | | 1,145 | | 836 | | 908 | | 1,053 | | 1,081 | | 1,087 | | 1,081 |
| July 2019 – June 2020 | | 1,217 | | 1,204 | | 1,150 | | 1,233 | | 1,264 | | 1,381 | | 994 | | 1,112 | | 1,202 | | 1,184 | | 1,138 | | 1,172 |
| year-on-year (%) | | 109.15 | | 114.67 | | 110.36 | | 114.59 | | 121.42 | | 120.61 | | 118.90 | | 122.47 | | 114.15 | | 109.53 | | 104.69 | | 108.42 |
| **Proportion of All Non-**  **Hodgkin Lymphoma to**  **All Cancers** | |  | |  | |  | |  | |  | |  | |  | |  | |  | |  | |  | |  |
| July 2018 – June 2019 | | 0.05 | | 0.05 | | 0.05 | | 0.05 | | 0.05 | | 0.05 | | 0.05 | | 0.05 | | 0.05 | | 0.05 | | 0.05 | | 0.05 |
| July 2019 – June 2020 | | 0.05 | | 0.05 | | 0.05 | | 0.06 | | 0.06 | | 0.06 | | 0.05 | | 0.05 | | 0.06 | | 0.06 | | 0.06 | | 0.06 |
| Ratio of proportion | | 1.05 | | 1.15 | | 1.10 | | 1.15 | | 1.21 | | 1.17 | | 1.14 | | 1.17 | | 1.15 | | 1.14 | | 1.10 | | 1.14 |
| **Chemotherapy** | |  | |  | |  | |  | |  | |  | |  | |  | |  | |  | |  | |  |
| July 2018 – June 2019 | | 877 | | 790 | | 828 | | 865 | | 829 | | 919 | | 648 | | 717 | | 834 | | 856 | | 875 | | 836 |
| July 2019 – June 2020 | | 958 | | 967 | | 930 | | 979 | | 1,028 | | 1,117 | | 803 | | 882 | | 975 | | 948 | | 931 | | 951 |
| year-on-year (%) | | 109.24 | | 122.41 | | 112.32 | | 113.18 | | 124.00 | | 121.55 | | 123.92 | | 123.01 | | 116.91 | | 110.75 | | 106.40 | | 113.76 |
| **Liver Cancer** | |  | |  | |  | |  | |  | |  | |  | |  | |  | |  | |  | |  |
| **All** | |  | |  | |  | |  | |  | |  | |  | |  | |  | |  | |  | |  |
| July 2018 – June 2019 | | 1,209 | | 1,215 | | 1,233 | | 1,231 | | 1,107 | | 1,288 | | 897 | | 1,125 | | 1,204 | | 1,179 | | 1,030 | | 1,106 |
| July 2019 – June 2020 | | 1,156 | | 1,102 | | 1,082 | | 1,096 | | 1,084 | | 1,128 | | 878 | | 1,093 | | 1,049 | | 1,058 | | 925 | | 1,016 |
| year-on-year (%) | | 95.62 | | 90.70 | | 87.75 | | 89.03 | | 97.92 | | 87.58 | | 97.88 | | 97.16 | | 87.13 | | 89.74 | | 89.81 | | 91.86 |
| **Proportion of All Liver**  **Cancer to All Cancers** | |  | |  | |  | |  | |  | |  | |  | |  | |  | |  | |  | |  |
| July 2018 – June 2019 | | 0.06 | | 0.05 | | 0.06 | | 0.06 | | 0.05 | | 0.06 | | 0.05 | | 0.06 | | 0.05 | | 0.05 | | 0.05 | | 0.05 |
| July 2019 – June 2020 | | 0.05 | | 0.05 | | 0.05 | | 0.05 | | 0.05 | | 0.05 | | 0.05 | | 0.05 | | 0.05 | | 0.05 | | 0.05 | | 0.05 |
| Ratio of proportion | | 0.92 | | 0.91 | | 0.87 | | 0.90 | | 0.98 | | 0.85 | | 0.94 | | 0.93 | | 0.88 | | 0.93 | | 0.95 | | 0.97 |
| **Surgery** | |  | |  | |  | |  | |  | |  | |  | |  | |  | |  | |  | |  |
| July 2018 – June 2019 | | 119 | | 120 | | 117 | | 128 | | 108 | | 112 | | 76 | | 109 | | 136 | | 94 | | 104 | | 93 |
| July 2019 – June 2020 | | 113 | | 140 | | 124 | | 111 | | 119 | | 141 | | 81 | | 120 | | 117 | | 109 | | 103 | | 101 |
| year-on-year (%) | | 94.96 | | 116.67 | | 105.98 | | 86.72 | | 110.19 | | 125.89 | | 106.58 | | 110.09 | | 86.03 | | 115.96 | | 99.04 | | 108.60 |
| **Chemotherapy** | |  | |  | |  | |  | |  | |  | |  | |  | |  | |  | |  | |  |
| July 2018 – June 2019 | | 144 | | 167 | | 152 | | 163 | | 132 | | 190 | | 136 | | 161 | | 176 | | 176 | | 142 | | 145 |
| July 2019 – June 2020 | | 172 | | 133 | | 166 | | 169 | | 166 | | 149 | | 117 | | 137 | | 134 | | 157 | | 138 | | 156 |
| year-on-year (%) | | 119.44 | | 79.64 | | 109.21 | | 103.68 | | 125.76 | | 78.42 | | 86.03 | | 85.09 | | 76.14 | | 89.20 | | 97.18 | | 107.59 |
| **RFA** | |  | |  | |  | |  | |  | |  | |  | |  | |  | |  | |  | |  |
| July 2018 – June 2019 | | 164 | | 146 | | 155 | | 193 | | 132 | | 171 | | 111 | | 145 | | 159 | | 155 | | 137 | | 149 |
| July 2019 – June 2020 | | 151 | | 166 | | 157 | | 151 | | 145 | | 133 | | 113 | | 126 | | 161 | | 140 | | 124 | | 134 |
| year-on-year (%) | | 92.07 | | 113.70 | | 101.29 | | 78.24 | | 109.85 | | 77.78 | | 101.80 | | 86.90 | | 101.26 | | 90.32 | | 90.51 | | 89.93 |
| **TACE** | |  | |  | |  | |  | |  | |  | |  | |  | |  | |  | |  | |  |
| July 2018 – June 2019 | | 397 | | 364 | | 422 | | 404 | | 348 | | 386 | | 255 | | 356 | | 356 | | 367 | | 293 | | 332 |
| July 2019 – June 2020 | | 352 | | 305 | | 300 | | 300 | | 333 | | 319 | | 250 | | 352 | | 305 | | 318 | | 246 | | 319 |
| year-on-year (%) | | 88.66 | | 83.79 | | 71.09 | | 74.26 | | 95.69 | | 82.64 | | 98.04 | | 98.88 | | 85.67 | | 86.65 | | 83.96 | | 96.08 |
| **Prostate Cancer** | |  | |  | |  | |  | |  | |  | |  | |  | |  | |  | |  | |  |
| **All** | |  | |  | |  | |  | |  | |  | |  | |  | |  | |  | |  | |  |
| July 2018 – June 2019 | | 684 | | 700 | | 622 | | 689 | | 678 | | 715 | | 579 | | 650 | | 732 | | 683 | | 645 | | 704 |
| July 2019 – June 2020 | | 754 | | 709 | | 686 | | 701 | | 721 | | 781 | | 626 | | 753 | | 736 | | 709 | | 598 | | 700 |
| year-on-year (%) | | 110.23 | | 101.29 | | 110.29 | | 101.74 | | 106.34 | | 109.23 | | 108.12 | | 115.85 | | 100.55 | | 103.81 | | 92.71 | | 99.43 |
| **Proportion of All**  **Prostate Cancer to All**  **Cancers** | |  | |  | |  | |  | |  | |  | |  | |  | |  | |  | |  | |  |
| July 2018 – June 2019 | | 0.03 | | 0.03 | | 0.03 | | 0.03 | | 0.03 | | 0.03 | | 0.03 | | 0.03 | | 0.03 | | 0.03 | | 0.03 | | 0.03 |
| July 2019 – June 2020 | | 0.03 | | 0.03 | | 0.03 | | 0.03 | | 0.03 | | 0.03 | | 0.03 | | 0.04 | | 0.03 | | 0.03 | | 0.03 | | 0.03 |
| Ratio of proportion | | 1.06 | | 1.02 | | 1.10 | | 1.02 | | 1.06 | | 1.06 | | 1.04 | | 1.10 | | 1.01 | | 1.08 | | 0.98 | | 1.05 |
| **Surgery** | |  | |  | |  | |  | |  | |  | |  | |  | |  | |  | |  | |  |
| July 2018 – June 2019 | | 238 | | 244 | | 214 | | 228 | | 234 | | 234 | | 193 | | 251 | | 278 | | 221 | | 204 | | 262 |
| July 2019 – June 2020 | | 270 | | 254 | | 255 | | 245 | | 251 | | 298 | | 205 | | 311 | | 290 | | 275 | | 244 | | 278 |
| year-on-year (%) | |  | |  | |  | |  | |  | |  | |  | |  | |  | |  | |  | |  |
| **TUR-P** | |  | |  | |  | |  | |  | |  | |  | |  | |  | |  | |  | |  |
| July 2018 – June 2019 | | 282 | | 274 | | 236 | | 326 | | 349 | | 294 | | 252 | | 298 | | 315 | | 267 | | 247 | | 271 |
| July 2019 – June 2020 | | 299 | | 289 | | 260 | | 261 | | 312 | | 311 | | 237 | | 305 | | 286 | | 240 | | 181 | | 221 |
| year-on-year (%) | | 106.03 | | 105.47 | | 110.17 | | 80.06 | | 89.40 | | 105.78 | | 94.05 | | 102.35 | | 90.79 | | 89.89 | | 73.28 | | 81.55 |
| **Chemotherapy** | |  | |  | |  | |  | |  | |  | |  | |  | |  | |  | |  | |  |
| July 2018 – June 2019 | | 174 | | 153 | | 158 | | 197 | | 172 | | 193 | | 136 | | 162 | | 182 | | 193 | | 172 | | 165 |
| July 2019 – June 2020 | | 196 | | 157 | | 176 | | 165 | | 177 | | 174 | | 174 | | 158 | | 175 | | 179 | | 153 | | 181 |
| year-on-year (%) | | 112.64 | | 102.61 | | 111.39 | | 83.76 | | 102.91 | | 90.16 | | 127.94 | | 97.53 | | 96.15 | | 92.75 | | 88.95 | | 109.70 |
| **Esophageal Cancer** | |  | |  | |  | |  | |  | |  | |  | |  | |  | |  | |  | |  |
| **All** | |  | |  | |  | |  | |  | |  | |  | |  | |  | |  | |  | |  |
| July 2018 – June 2019 | | 738 | | 725 | | 711 | | 715 | | 682 | | 736 | | 641 | | 672 | | 654 | | 766 | | 665 | | 688 |
| July 2019 – June 2020 | | 746 | | 724 | | 685 | | 694 | | 670 | | 724 | | 620 | | 674 | | 699 | | 620 | | 592 | | 620 |
| year-on-year (%) | | 101.08 | | 99.86 | | 96.34 | | 97.06 | | 98.24 | | 98.37 | | 96.72 | | 100.30 | | 106.88 | | 80.94 | | 89.02 | | 90.12 |
| **Proportion of All**  **Esophageal Cancer to**  **All Cancers** | |  | |  | |  | |  | |  | |  | |  | |  | |  | |  | |  | |  |
| July 2018 – June 2019 | | 0.03 | | 0.03 | | 0.03 | | 0.03 | | 0.03 | | 0.03 | | 0.04 | | 0.03 | | 0.03 | | 0.03 | | 0.03 | | 0.03 |
| July 2019 – June 2020 | | 0.03 | | 0.03 | | 0.03 | | 0.03 | | 0.03 | | 0.03 | | 0.03 | | 0.03 | | 0.03 | | 0.03 | | 0.03 | | 0.03 |
| Ratio of proportion | | 0.97 | | 1.01 | | 0.96 | | 0.98 | | 0.98 | | 0.95 | | 0.93 | | 0.96 | | 1.08 | | 0.84 | | 0.94 | | 0.95 |
| **Surgery** | |  | |  | |  | |  | |  | |  | |  | |  | |  | |  | |  | |  |
| July 2018 – June 2019 | | 42 | | 46 | | 50 | | 60 | | 53 | | 50 | | 46 | | 37 | | 57 | | 42 | | 60 | | 42 |
| July 2019 – June 2020 | | 59 | | 44 | | 41 | | 60 | | 41 | | 51 | | 35 | | 49 | | 54 | | 40 | | 46 | | 47 |
| year-on-year (%) | | 140.48 | | 95.65 | | 82.00 | | 100.00 | | 77.36 | | 102.00 | | 76.09 | | 132.43 | | 94.74 | | 95.24 | | 76.67 | | 111.90 |
| **Endoscopic resection** | |  | |  | |  | |  | |  | |  | |  | |  | |  | |  | |  | |  |
| July 2018 – June 2019 | | 120 | | 106 | | 118 | | 108 | | 102 | | 121 | | 90 | | 113 | | 88 | | 124 | | 80 | | 97 |
| July 2019 – June 2020 | | 119 | | 109 | | 120 | | 105 | | 105 | | 125 | | 94 | | 119 | | 125 | | 112 | | 79 | | 82 |
| year-on-year (%) | | 99.17 | | 102.83 | | 101.69 | | 97.22 | | 102.94 | | 103.31 | | 104.44 | | 105.31 | | 142.05 | | 90.32 | | 98.75 | | 84.54 |
| **Chemotherapy** | |  | |  | |  | |  | |  | |  | |  | |  | |  | |  | |  | |  |
| July 2018 – June 2019 | | 333 | | 324 | | 291 | | 306 | | 299 | | 319 | | 307 | | 298 | | 310 | | 346 | | 296 | | 326 |
| July 2019 – June 2020 | | 322 | | 317 | | 292 | | 306 | | 316 | | 326 | | 303 | | 289 | | 304 | | 263 | | 290 | | 286 |
| year-on-year (%) | | 96.70 | | 97.84 | | 100.34 | | 100.00 | | 105.69 | | 102.19 | | 98.70 | | 96.98 | | 98.06 | | 76.01 | | 97.97 | | 87.73 |
| **Uterine Cancer** | |  | |  | |  | |  | |  | |  | |  | |  | |  | |  | |  | |  |
| **All** | |  | |  | |  | |  | |  | |  | |  | |  | |  | |  | |  | |  |
| July 2018 – June 2019 | | 631 | | 670 | | 625 | | 614 | | 608 | | 644 | | 554 | | 580 | | 642 | | 610 | | 607 | | 647 |
| July 2019 – June 2020 | | 719 | | 711 | | 619 | | 669 | | 655 | | 629 | | 544 | | 596 | | 629 | | 627 | | 601 | | 600 |
| year-on-year (%) | | 113.95 | | 106.12 | | 99.04 | | 108.96 | | 107.73 | | 97.67 | | 98.19 | | 102.76 | | 97.98 | | 102.79 | | 99.01 | | 92.74 |
| **Proportion of All Uterine**  **Cancer to All Cancers** | |  | |  | |  | |  | |  | |  | |  | |  | |  | |  | |  | |  |
| July 2018 – June 2019 | | 0.03 | | 0.03 | | 0.03 | | 0.03 | | 0.03 | | 0.03 | | 0.03 | | 0.03 | | 0.03 | | 0.03 | | 0.03 | | 0.03 |
| July 2019 – June 2020 | | 0.03 | | 0.03 | | 0.03 | | 0.03 | | 0.03 | | 0.03 | | 0.03 | | 0.03 | | 0.03 | | 0.03 | | 0.03 | | 0.03 |
| Ratio of proportion | | 1.10 | | 1.07 | | 0.98 | | 1.10 | | 1.08 | | 0.95 | | 0.95 | | 0.98 | | 0.99 | | 1.07 | | 1.04 | | 0.98 |
| **Surgery** | |  | |  | |  | |  | |  | |  | |  | |  | |  | |  | |  | |  |
| July 2018 – June 2019 | | 190 | | 162 | | 146 | | 176 | | 162 | | 178 | | 141 | | 193 | | 171 | | 199 | | 160 | | 198 |
| July 2019 – June 2020 | | 231 | | 206 | | 175 | | 179 | | 185 | | 197 | | 127 | | 158 | | 188 | | 213 | | 173 | | 155 |
| year-on-year (%) | | 121.58 | | 127.16 | | 119.86 | | 101.70 | | 114.20 | | 110.67 | | 90.07 | | 81.87 | | 109.94 | | 107.04 | | 108.13 | | 78.28 |
| **Chemotherapy** | |  | |  | |  | |  | |  | |  | |  | |  | |  | |  | |  | |  |
| July 2018 – June 2019 | | 343 | | 367 | | 350 | | 330 | | 335 | | 352 | | 313 | | 293 | | 358 | | 312 | | 324 | | 336 |
| July 2019 – June 2020 | | 356 | | 371 | | 331 | | 369 | | 362 | | 306 | | 324 | | 331 | | 315 | | 316 | | 323 | | 314 |
| year-on-year (%) | | 103.79 | | 101.09 | | 94.57 | | 111.82 | | 108.06 | | 86.93 | | 103.51 | | 112.97 | | 87.99 | | 101.28 | | 99.69 | | 93.45 |
| **Gallbladder & Biliary tract Cancer** | |  | |  | |  | |  | |  | |  | |  | |  | |  | |  | |  | |  |
| **All** | |  | |  | |  | |  | |  | |  | |  | |  | |  | |  | |  | |  |
| July 2018 – June 2019 | | 571 | | 636 | | 543 | | 559 | | 546 | | 666 | | 467 | | 509 | | 575 | | 558 | | 538 | | 589 |
| July 2019 – June 2020 | | 610 | | 565 | | 583 | | 597 | | 531 | | 638 | | 491 | | 570 | | 540 | | 565 | | 539 | | 580 |
| year-on-year (%) | | 106.83 | | 88.84 | | 107.37 | | 106.80 | | 97.25 | | 95.80 | | 105.14 | | 111.98 | | 93.91 | | 101.25 | | 100.19 | | 98.47 |
| **Proportion of**  **Gallbladder & Biliary**  **tract Cancer to All   Cancers** | |  | |  | |  | |  | |  | |  | |  | |  | |  | |  | |  | |  |
| July 2018 – June 2019 | | 0.03 | | 0.03 | | 0.03 | | 0.03 | | 0.03 | | 0.03 | | 0.03 | | 0.03 | | 0.03 | | 0.03 | | 0.03 | | 0.03 |
| July 2019 – June 2020 | | 0.03 | | 0.03 | | 0.03 | | 0.03 | | 0.02 | | 0.03 | | 0.03 | | 0.03 | | 0.02 | | 0.03 | | 0.03 | | 0.03 |
| Ratio of proportion | | 1.03 | | 0.89 | | 1.07 | | 1.07 | | 0.97 | | 0.93 | | 1.01 | | 1.07 | | 0.95 | | 1.05 | | 1.06 | | 1.04 |
| **Surgery** | |  | |  | |  | |  | |  | |  | |  | |  | |  | |  | |  | |  |
| July 2018 – June 2019 | | 26 | | 45 | | 26 | | 27 | | 27 | | 44 | | 34 | | 34 | | 39 | | 30 | | 45 | | 34 |
| July 2019 – June 2020 | | 38 | | 43 | | 27 | | 49 | | 27 | | 30 | | 22 | | 30 | | 24 | | 32 | | 24 | | 36 |
| year-on-year (%) | | 146.15 | | 95.56 | | 103.85 | | 181.48 | | 100.00 | | 68.18 | | 64.71 | | 88.24 | | 61.54 | | 106.67 | | 53.33 | | 105.88 |
| **Chemotherapy** | |  | |  | |  | |  | |  | |  | |  | |  | |  | |  | |  | |  |
| July 2018 – June 2019 | | 93 | | 121 | | 91 | | 95 | | 91 | | 100 | | 62 | | 61 | | 73 | | 71 | | 87 | | 88 |
| July 2019 – June 2020 | | 105 | | 85 | | 85 | | 84 | | 89 | | 80 | | 75 | | 94 | | 66 | | 77 | | 80 | | 83 |
| year-on-year (%) | | 112.90 | | 70.25 | | 93.41 | | 88.42 | | 97.80 | | 80.00 | | 120.97 | | 154.10 | | 90.41 | | 108.45 | | 91.95 | | 94.32 |

Abbreviations; TUR-BT: Transurethral resection of bladder tumor, RFA: Radiofrequency ablation, TACE: Transcatheter arterial chemoembolization, TUR-P: Transurethral resection of prostate

*: The ratio of proportions = proportion of admission cases [2019/07-2020/06] / proportion of admission cases [2018/07-2019/06]

where

proportion of admission cases [2019/7-2020/6] = number of admission cases with each diagnosis of cancer per month between July 2019 and June 2020 / number of admission cases with all cancers per month between July 2019 and June 2020

and where

proportion of admission cases [2018/7-2019/6] = number of admission cases with each diagnosis of cancer per month between July 2018 and June 2019 / number of admission cases with all cancers per month between July 2018 and June 2019.
